# Fibroblast Growth Factor-23 and Risk of Cardiovascular Diseases: a Mendelian Randomisation study

**DOI:** 10.1101/2022.04.27.22273667

**Authors:** Killian Donovan, William G. Herrington, Guillaume Paré, Marie Pigeyre, Richard Haynes, Rebecca Sardell, Adam S. Butterworth, Lasse Folkersen, Stefan Gustafsson, Qin Wang, Colin Baigent, Anders Mälarstig, Michael Holmes, Natalie Staplin

## Abstract

Fibroblast growth factor 23 (FGF-23) is associated with a range of cardiovascular and non-cardiovascular diseases in conventional epidemiological studies, but substantial residual confounding may exist. Mendelian randomisation approaches can help control for such confounding. SCALLOP consortium data on 19,195 participants were used to generate an FGF-23 genetic score. Data from 337,448 UK Biobank participants were used to estimate associations between higher genetically-predicted FGF-23 concentration and the odds of any atherosclerotic cardiovascular disease (n=26,266 events), of any non-atherosclerotic cardiovascular disease (n=12,652), and of non-cardiovascular diseases previously linked to FGF-23. Measurements of carotid intima-media thickness (CIMT) and left ventricular mass (LVM) were available in a subset. Associations with cardiovascular outcomes were also tested in three large case-control consortia: CARDIOGRAMplusC4D (coronary artery disease, n=181,249 cases), MEGASTROKE (stroke, n=34,217), and HERMES (heart failure, n=47,309). We identified 34 independent variants for circulating FGF-23 which formed a validated genetic score. There were no associations between genetically-predicted FGF-23 and any of the cardiovascular or non-cardiovascular outcomes. In UK Biobank, the odds ratio for any atherosclerotic cardiovascular disease per 1-SD higher genetically-predicted logFGF-23 was 1.03 (95% confidence interval [CI] 0.98-1.08), and for any non-atherosclerotic cardiovascular disease was 1.01 (0.94-1.09). The odds ratios in the case-control consortia were 1.00 (0.97-1.03) for coronary artery disease, 1.01 (0.95-1.07) for stroke, and 1.00 (0.95-1.05) for heart failure. In those with imaging, logFGF-23 was not associated with CIMT or LVM index. This suggests that previously reported observational associations of FGF-23 with risk of atherosclerotic and non-atherosclerotic cardiovascular diseases are unlikely to be causal.

## Introduction

Higher risk of cardiovascular disease emerges early in the development of chronic kidney disease (CKD), and risk progressively increases as kidney function declines.^1,2^ Arterial disease in advanced CKD exhibits non-atheromatous non-calcified arterial stiffening, intimal atherosclerotic lesions, and heavy medial calcification.^3^ Correspondingly, CKD is associated with both structural and coronary heart disease. Perhaps about one-half of this risk is explained by the effects of CKD on blood pressure.^3^ Non-traditional risk factors associated with dysregulated phosphate and calcium homeostasis may also be important.^4-6^

Fibroblast growth factor-23 (FGF-23) is a hormonal promoter of urinary phosphate excretion, increasing in blood concentration in early CKD.^7^ The actions of FGF-23 are generally limited to tissues where the co-receptor Klotho is expressed, and particularly in the renal tubules where it downregulates sodium-phosphate co-transporters.^8,9^ Animal studies suggest direct (i.e. Klotho-independent effect) cardiotoxicity,^10^ offering a biological explanation for observed associations between FGF-23 and left ventricular hypertrophy^11,12^ and leading to hypotheses that FGF23 should be considered not just as a marker for cardiovascular disease, but also as a causal contributory factor.^13^ A positive association between FGF-23 and carotid intima-media thickness (CIMT) has also been reported.^14^ Meta-analysis of conventional epidemiological studies has found independent associations between higher circulating FGF-23 concentration with higher risk of atherosclerotic cardiovascular diseases (i.e. myocardial infarction and stroke) and heart failure.^15^ However, substantial uncertainty about causality remains as these associations do not exhibit a clear “exposure-response” relationship and are non-specific: positive associations between FGF-23 and risks of infection,^16^ fractures,^17^ acute kidney injury^18^ and all-cause mortality^19^ are also reported. Residual confounding therefore remains a possible explanation for these FGF-23 associations.

Naturally occurring genetic variants (single nucleotide polymorphisms [SNPs]) associated with biological traits are allocated randomly at conception and can be used as instruments in genetic epidemiological analyses. This Mendelian randomisation (MR) approach can avoid some of the limitations inherent to conventional observational studies.^20-22^ MR has a particular advantage when studying FGF-23, as completely controlling for confounding by kidney function in conventional studies is challenging by virtue of both serum creatinine’s natural within-person variability, and the inherent imprecision when using it to estimate kidney function in those with normal or early CKD.^23^ Previously reported MR studies of FGF-23 have been limited by low power as the genetic variants used explain only ∼3% of the variation in FGF-23, and they have not explored the breakdown of associations with atherosclerotic versus non-atherosclerotic phenotypes.^24-27^ We aimed to derive a more powerful genetic score for FGF-23 from a large international collaboration’s genotypic and proteomic data, and then use it to estimate associations between lifelong genetically-predicted differences in circulating FGF-23 with risk of cardiovascular diseases in the UK Biobank cohort, and in the CARDIOGRAMplusC4D, MEGASTROKE, and HERMES-HF case-control consortia. We considered genetic associations for atherosclerotic and non-atherosclerotic cardiovascular phenotypes separately, as well as a range of non-cardiovascular diseases identified in non-genetic epidemiological studies.

## Methods

### Study populations and data

Table 1 summarizes the study design and the different study populations used to derive and validate the novel FGF-23 genetic score, and then test associations with clinical outcomes and measurements. The score was derived and validated in cohorts with a low CKD prevalence in order to reduce the risk of identifying pleiotropic variants associated with FGF-23 only through their association with kidney disease.

**Table 1:**
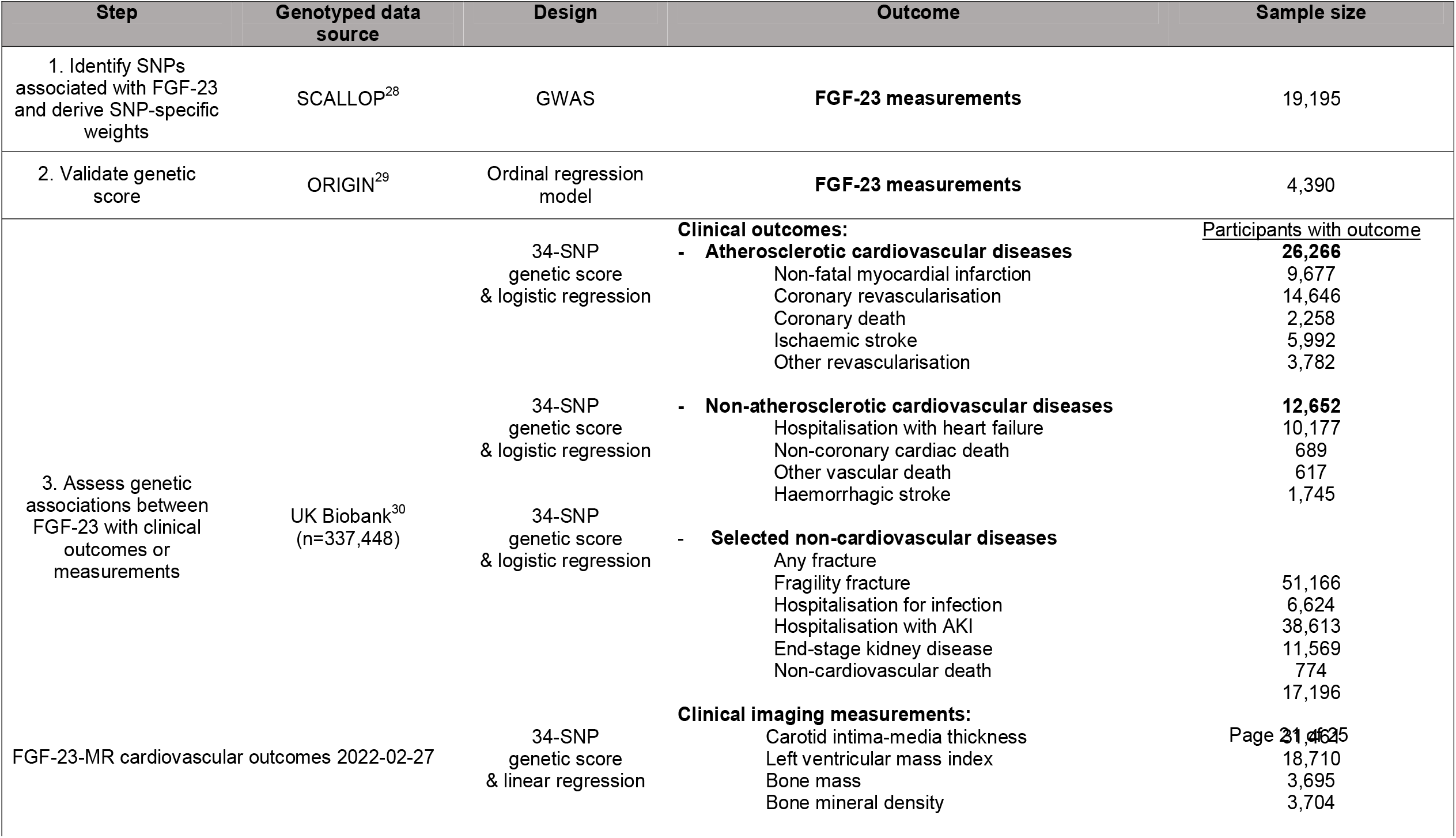
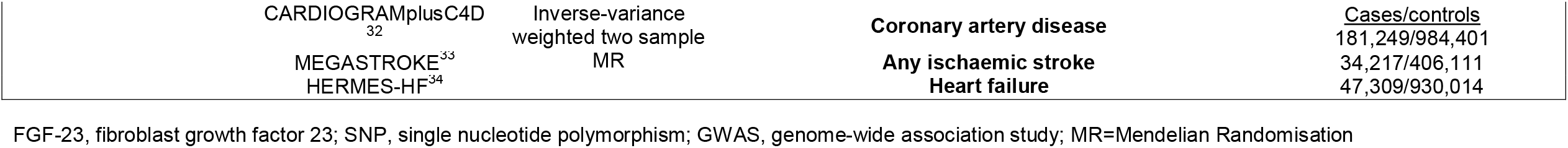
Study design and sources of data.

SCALLOP Consortium data from 19,195 individuals were used to identify variants associated with FGF-23 for a novel genetic score. SCALLOP is a collaboration of genotyped cohorts with proteomic measurements using the multiplex immunoassay Olink platform.^28^ The assay uses two antibodies which separately bind at FGF-23’s N-terminus and C-terminus, analogous to intact FGF-23 assays.

Once derived, the genetic score was validated in an independent cohort of 4,390 genotyped individuals from the ORIGIN trial in people with dysglycaemia whose blood had also been assayed for FGF-23 using a Luminex platform ^29^ (Myriad-RBM).

Associations between the FGF-23 genetic score and risk of clinical outcomes and measurements were first assessed in UK Biobank, a prospective genotyped cohort of 502,650 UK adults aged 40-69 years recruited between 2006 and 2010.^30^ UK Biobank data include self-completed touch-screen questionnaires, computer-assisted interviews, physical and functional measurements, and biochemical assays. Genome-wide genotyping was performed using the Affymetrix UK BiLEVE Axiom array and the Affymetrix UK Biobank Axiom array, with imputation with IMPUTE4 using the Haplotype Reference Consortium and the UK 10K and 1000 Genomes phase 3 reference panels.^31^ UK Biobank has been linked to routinely collected UK mortality and hospital admission data from which a range of clinical outcomes can be ascertained. To ensure comparability with the SCALLOP derivation cohort (> 90% of European ancestry), the UKB analyses included unrelated white British participants with available genetic data meeting quality control standards (n=335,536). Participants were excluded if they had withdrawn their data from the UK Biobank (n=88). A subset of these participants has also undergone carotid ultrasound imaging (n=31,461 were included in the present study), cardiac magnetic resonance imaging (n=18,734), and DEXA scanning of bone mass (n=3,695).

Associations between FGF-23 with risk of specific cardiovascular outcomes were also assessed in 3 large case-control consortia: CARDIOGRAMplusC4D,^32^ MEGASTROKE,^33^ and HERMES.^34^ These international case-control consortia comprise genomic and clinical outcome data for individuals with coronary artery disease (181,249 cases/984,401 controls), ischaemic stroke (34,217 cases/406,111 controls), and heart failure (47,309 cases/930,014 controls), respectively.

### FGF-23 genetic instrument selection

Autosomal variants associated with FGF-23 at p<5×10^−6^ in SCALLOP Consortium data were clumped using PLINK^35^ to identify independent variants (r^2^<0.1; 1000 Genomes Phase 3 reference panel). A genetic score for individual participants was constructed with a weighted sum of the dosages of these SNPs, using the SCALLOP estimates as weights for each variant. Associations between this score and measured FGF-23 levels were confirmed in the ORIGIN cohort.^29^ Any *cis* variants within 100kb of the FGF-23 transcription start site were also selected as instruments for sensitivity analyses.

### Outcomes

Study outcomes were selected based on those previously observed conventional observational associations with FGF-23.^15^ The key cardiovascular outcomes in UK Biobank were “any atherosclerotic cardiovascular disease”, a composite of coronary death, non-fatal myocardial infarction, ischaemic stoke, or revascularization, and separately “any non-atherosclerotic cardiovascular disease”, a composite of non-coronary cardiac and other vascular death, hospitalisation with heart failure, or haemorrhagic stroke. Non-cardiovascular outcomes included any bone fracture, and the subset with a fragility fracture, hospitalisation for infection, hospitalisation with acute kidney injury, treated end-stage kidney disease, and any non-cardiovascular death (Supplemental Methods provide the list of diagnostic and procedural codes used in outcomes). In the UK Biobank subset of participants with magnetic resonance imaging, left ventricular mass (LVM) index was established using previously published algorithms.^36^ Mean and maximum CIMT were used among the subset with carotid ultrasound assessments. Android/gynoid bone mass and bone mineral density of lumbar vertebrae and the femoral neck were used as measurements of bone health.

### Statistical analyses

The genetic score was validated by regression of measured FGF-23 on the genetic score in the ORIGIN cohort, adjusted for age, sex and ethnicity. Ordinal regression was used due to a high proportion (59%) of participants in ORIGIN with FGF-23 levels below the lower limit of detection of the assay. After completing validation of the genetic score and deriving outcome data, the power of our genetic instrument was approximated using the proportion of variance explained using the methods of Burgess et al,^37^ and also expressed as the minimum detectable true odds ratio per unit higher genetic score (OR) at 80% power and α=0.05.^38^ A one unit change in the genetic score was scaled to correspond to a 1-standard deviation (SD) change in genetically-predicted logFGF-23. Estimates of associations between binary clinical outcomes and the genetic score were ascertained from logistic regression models adjusted for age, sex and the first 40 genomic principal components. Linear regression models including these same covariates were used for continuous outcomes. Analyses of case-control consortia data employed a two-sample inverse variance weighted MR method with summary data for the 34 SNPs (SNP-logFGF23 associations from SCALLOP and SNP-outcome associations from relevant consortia). For the key analyses of cardiovascular and non-cardiovascular outcomes, a significance level of p<0.05 was used for all individual statistical tests. For the subsidiary assessments using imaging-based clinical measurements, and for sensitivity analyses a Bonferroni correction was applied to p values.

The key sensitivity analysis was to assess association for the variants within 100kb of the FGF-23 transcription start site (i.e. any *cis-*variants), as such variants are less likely to have unidentified pleiotropic effects.^39^ We also performed a sensitivity analysis excluding any potentially pleiotropic variants identified to modify cardiovascular risk factors other than FGF-23 (e.g. blood pressure, blood lipids, propensity to smoke cigarettes or develop diabetes mellitus). Such variants were identified from published associations or UK Biobank using the PhenoScanner database^40,41^ and significance threshold of p<0.01. Any effect of any linkage disequilibrium (LD) between selected variants was assessed in a sensitivity analysis using a genetic score derived with a more restrictive LD threshold (r^2^<0.001 [1000 Genomes Phase 3 Release]). Effect estimates from different MR approaches were compared to assess validity of the instrumental variable assumptions (relevance, independence and exclusion restriction).^42-44^ MR analyses were performed in SAS version 9.4 (SAS Institute, Cary NY, USA) and R v3.6.2.

## Results

### Genetic instrument derivation, validation and power

Thirty-four independent (r^2^<0.1) variants associated at p<5×10^−6^ with circulating FGF-23 concentration were identified from SCALLOP Consortium data (see Supplemental Table 1 for details of each SNP). Of these variants, three were found to be associated with other cardiovascular risk factors other than FGF-23 (see Supplemental Table 2), and two independent *cis-*variants were identified (rs6489536 [FGF-23 increasing allele: C], rs7955866 [FGF-23 increasing allele: G]; r^2^=0.08; Supplemental Tables 2&3) were identified. The 34 SNPs collectively accounted for 6.3% of the variance in log-transformed FGF-23 (Supplemental Table 4), and the two *cis*-variants accounted for 0.4%. A higher genetic score was associated with higher measured FGF-23 in ORIGIN, validating the novel score (Supplemental Table 5). The genetic score and number of atherosclerotic and non-atherosclerotic cardiovascular outcomes in the included UK Biobank population provided 80% power at α=0.05 to detect a 1.08 and 1.11 minimum OR per 1-SD higher logFGF-23, respectively (Supplemental Table 4).

### UK Biobank population characteristics

Median age among the 337,448 included UK Biobank participants was 58 (Q1-Q3: 51-63) years, 181,216 (53.7%) were female, 33,971 (10.1%) were current smokers, and 16,218 (4.8%) reported a history of diabetes mellitus. Mean (SD) body mass index was 27.4 (4.7) kg/m^2^ and median estimated glomerular filtration rate (eGFR) was 91 (81-100) mL/min/1.73m^2^. There were no important differences in age, sex, lifestyle factors, anthropometric measurements, blood pressure, prevalence of diabetes mellitus, glycosylated haemoglobin levels, eGFR, and markers of calcium/phosphate homeostasis across fifths of the FGF-23 genetic score (Table 2).

**Table 2:** Baseline characteristics of included UK Biobank participant by fifths of the FGF-23 genetic score.

|  | All participants<br>(n=337,448) | Bottom fifth of score<br>(n=67,489) | Middle fifth of score<br>(n=67,489) | Top fifth of score<br>(n=67,486) | Difference<br>between top and<br>bottom fifth |
| --- | --- | --- | --- | --- | --- |
| **Age , years** | **58 (51 - 63)** | 58 (51 - 63) | 58 (51 - 63) | 58 (51 - 63) | **0.0** |
| **Female** | **181216 (53.7)** | 36288 (53.8) | 36266 (53.7) | 36310 (53.8) | **0.0%** |
| **Current smoker** | **33971 (10.1)** | 6694 (9.9) | 6870 (10.2) | 6753 (10.0) | **0.1%** |
| **Body mass index (kg/m<sup>2</sup>)** | **27.4 (4.7)** | 27.4 (4.8) | 27.4 (4.8) | 27.4 (4.8) | **0.0** |
| **Waist-to-hip ratio** | **0.87 (0.09)** | 0.87 (0.09) | 0.87 (0.09) | 0.87 (0.09) | **0.0** |
| **Diabetes mellitus**<br>**(self-report)** | **16218 (4.8)** | 3332 (4.9) | 3272 (4.8) | 3214 (4.8) | **0.1%** |
| HbA1c (mmol/mol) | **35.1 (32.7 – 37.7)** | 35.1 (32.7 – 37.8) | 35.2 (32.7 – 37.7) | 35.2 (32.8 – 37.8) | **0.1** |
| **Blood pressure** |  |  |  |  |  |
| Systolic (mmHg) | **138.3 (18.6)** | 138.2 (18.6) | 138.3 (18.5) | 138.3 (18.6) | **0.1** |
| Diastolic (mmHg) | **82.3 (10.1)** | 82.3 (10.1) | 82.3 (10.1) | 82.3 (10.1) | **0.0** |
| Antihypertensive use | **78433 (23.2%)** | 15678 (23.2%) | 15790 (23.4%) | 15554 (23.0%) | **0.2%** |
| **Kidney function** |  |  |  |  |  |
| eGFR (ml/min/1.73m<sup>2</sup>) | **90.8 (81.1 – 100.0)** | 90.8 (81.2 – 99.9) | 90.9 (81.3 – 100.0) | 90.1 (81.1 – 100.0) | **0.7** |
| eGFR <45 or on RRT | **1994 (0.6)** | 392 (0.6) | 403 (0.6) | 421 (0.6) | **0.0%** |
| eGFR ≥45 | **319362 (94.6)** | 63950 (94.8) | 63869 (94.6) | 63709 (94.4) | **0.4%** |
| eGFR missing | **16092 (4.8)** | 3147 (4.7) | 3217 (4.8) | 3356 (5.0) | **0.3%** |
| Urinary uACR >3 mg/mmol | **16463 (4.9)** | 3310 (4.9) | 3277 (4.9) | 3281 (4.9) | **0.0%** |
| **CholesterolCholesterol** |  |  |  |  |  |
| LDL cholesterol (mmol/L) | **3.5 (3.0 – 4.1)** | 3.5 (3.0 – 4.1) | 3.5 (3.0 – 4.1) | 3.5 (3.0 – 4.1) | **0.0** |
| HDL cholesterol (mmol/L) | **1.4 (1.2 – 1.7)** | 1.4 (1.2 – 1.7) | 1.4 (1.2 – 1.7) | 1.4 (1.2 – 1.7) | **0.0** |
| **Markers of calcium/phosphate homeostasis** |  |  |  |  |  |
| Calcium (mmol/L) | **2.38 (0.09)** | 2.38 (0.09) | 2.38 (0.09) | 2.38 (0.09) | **0.00** |
| Phosphate (mmol/L) | **1.16 (0.16)** | 1.15 (0.16) | 1.16 (0.16) | 1.16 (0.16) | **0.01** |
| Vitamin D (ng/mL) | **49.8 (21.0)** | 49.4 (20.9) | 49.9 (20.9) | 50.1 (21.1) | **0.7** |

Data are mean (SD), n (%) or median (Q1-Q3). FGF-23, Fibroblast growth factor 23; HbA1c, glycosylated haemoglobin; eGFR, estimated glomerular filtration rate; RRT, renal replacement therapy; uACR, urinary albumin:creatinine ratio; LDL, low-density lipoprotein cholesterol; HDL, high-density lipoprotein cholesterol; Blood pressure is the mean of two measurements. eGFR calculated by CKD-EPI combined creatinine + cystatin C equation (Inker et al. N Engl J Med 2012; 367:20-29, 2012)

### FGF-23 and risk of atherosclerotic cardiovascular disease

26,266 (7.8%) of the UK Biobank participants had atherosclerotic cardiovascular disease. There was no association between genetically-predicted FGF23 with the composite of any atherosclerotic cardiovascular disease (OR per 1-SD higher logFGF-23=1.03, 95% confidence interval [95%CI] 0.98-1.08). Nor were there any significant associations with any of its constituent components: non-fatal myocardial infarction (1.07, 0.99-1.16; 9,677 outcomes), coronary death (1.14, 0.97-1.34; 2,258 outcomes), ischaemic stroke (1.04, 0.94– 1.15; 5,992 outcomes), coronary revascularisation (1.01, 0.95–1.08; 14,646 outcomes) or other revascularisation (0.99, 0.87–1.13; 3,782 outcomes; Figure 1).

**Figure 1:**
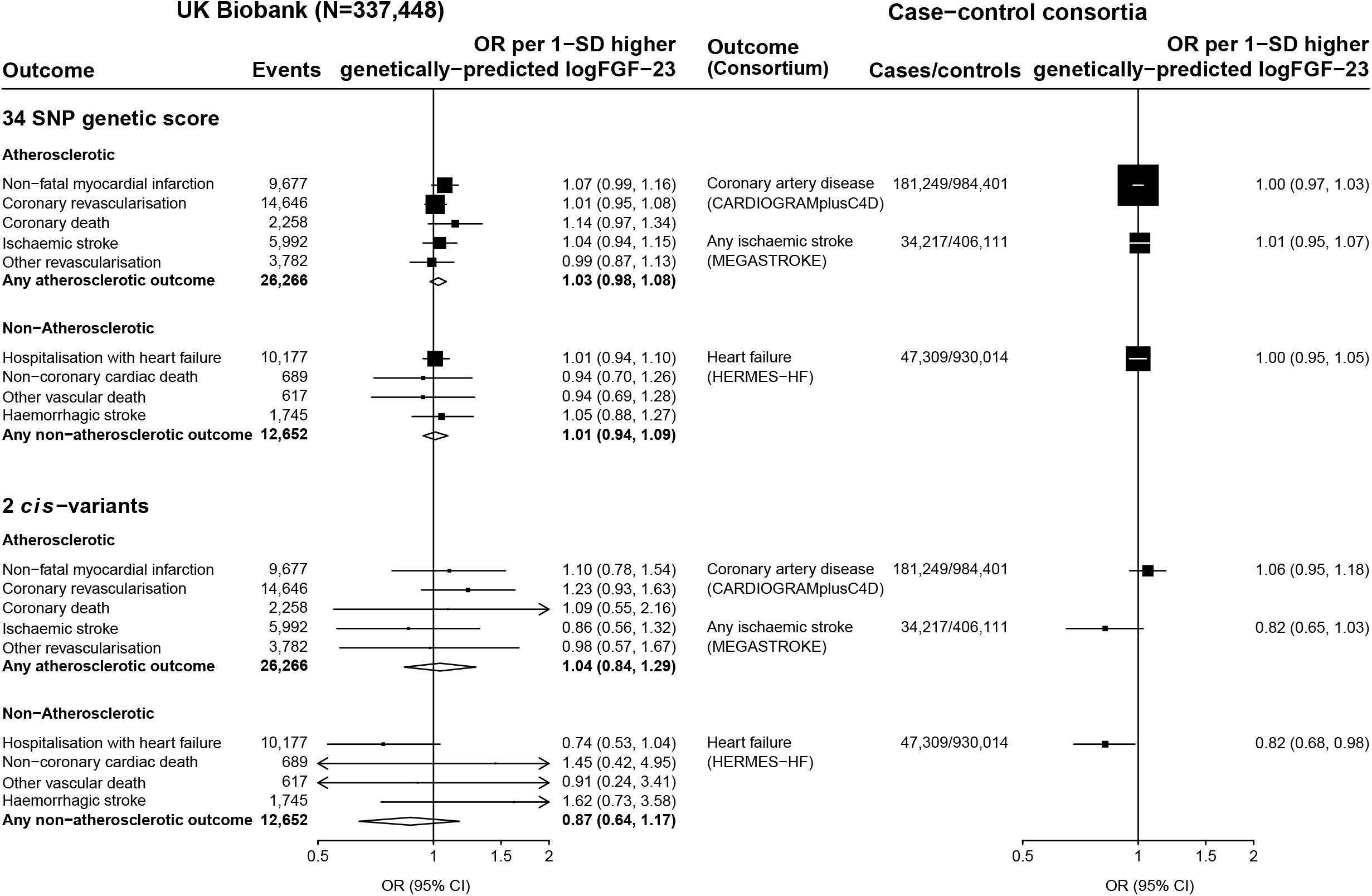
Associations between genetically predicted FGF-23 with risk of cardiovascular outcomes. FGF-23, Fibroblast growth factor 23; SNP, single nucleotide polymorphism; OR, odds ratio; SD, standard deviation; 95% CI, 95% confidence interval.

Analyses of 1.17 million participants in CARDIoGRAMplusC4D found no significant association between genetically predicted FGF-23 and coronary artery disease (OR per 1-SD higher genetically predicted log[FGF-23] 1.00; 95%CI 0.97-1.03; 181,249 cases). In the European-ancestry subset of the MEGASTROKE consortium (440,328 participants), 30 of the FGF-23 SNPs were available (missing SNPs: rs11542063, rs117612483, rs117989952, rs189972262 with no available strong proxies). Inverse variance weighted MR also demonstrated no significant association between genetically-predicted FGF-23 with risk of ischaemic stroke (OR per 1-SD higher genetically predicted log[FGF-23] 1.01, 0.95–1.07; 34,217 cases: Figure 1).

Among the subset of 31,461 UK Biobank participants with carotid imaging, there was no significant association between genetically-predicted FGF-23 and CIMT. A 1-SD higher genetically predicted FGF23 was associated with a -1μm difference in mean CIMT (95% CI - 6 to 4μm) and a 0μm difference in maximum CIMT (−6 to 6μm: Table 3).

**Table 3:**
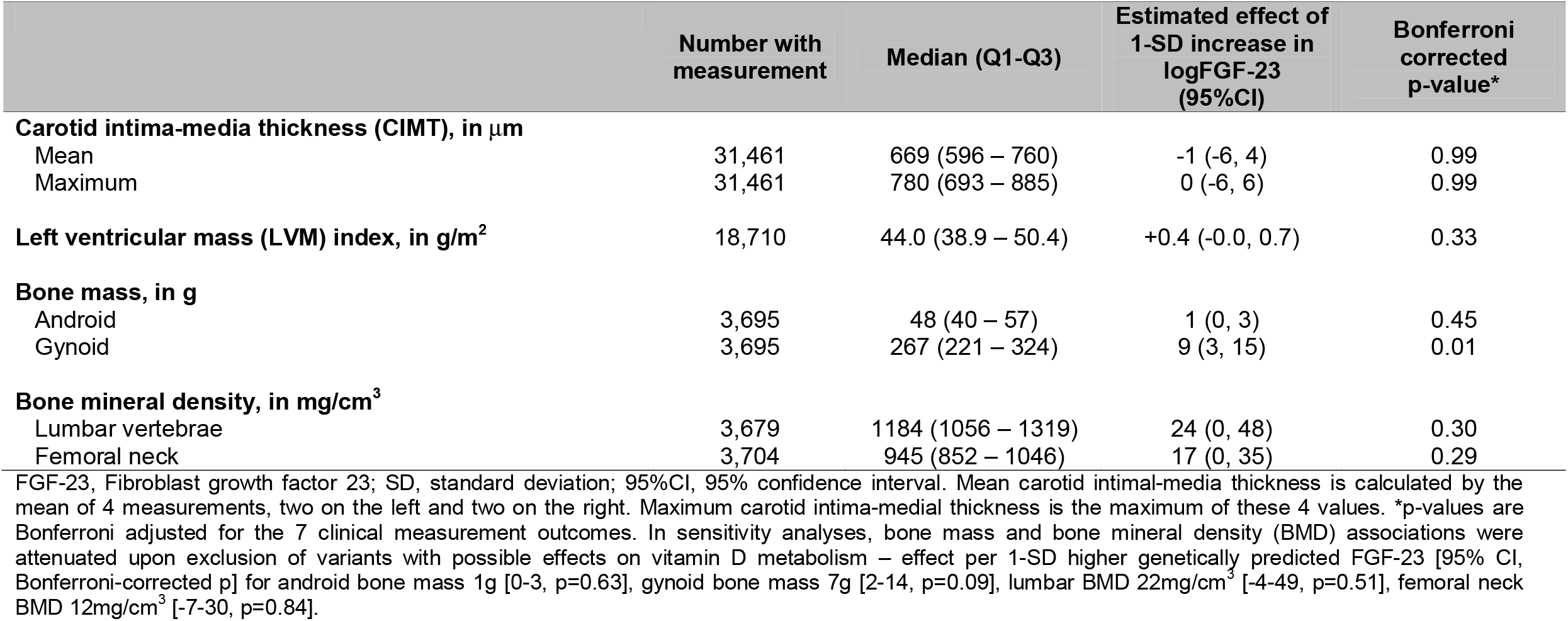
Association of genetically predicted FGF-23 with clinical measurements.

### FGF-23 and risk of non-atherosclerotic cardiovascular disease

12,652 (3.7%) UK Biobank participants had a non-atherosclerotic cardiovascular disease. There was no association between genetically-predicted FGF23 with any non-atherosclerotic cardiovascular disease (OR per 1-SD higher logFGF23=1.01, 0.94-1.09). This included no significant association with its constituent components: hospitalisation with heart failure (1.01, 0.94-1.10; 10,177 outcomes), non-coronary cardiac death (0.94, 0.70-1.26; 689 outcomes), non-cardiac vascular death (0.94, 0.69-1.28; 617 outcomes) or haemorrhagic stroke (1.05, 0.88-1.27; 1,745 outcomes; Figure 1).

Amongst 977,323 participants in HERMES-HF, 29 of the FGF-23 associated variants identified in SCALLOP were available (missing SNPs: rs11542063, rs117612483, rs117989952, rs189972262, rs75357988, with no available strong proxies). Inverse variance weighted MR using data for the remaining 29 SNPs found no significant association between genetically-predicted FGF-23 with heart failure (OR per 1-SD higher genetically predicted log[FGF-23] 1.00, 0.95-1.05; 47,309 cases; Figure 1).

Among the UK Biobank subset with cardiac magnetic resonance imaging, the median (Q1-Q3) LVM index (i.e. LVM per square metre of body surface area) was 44.0g/m^2^ (38.9–50.4). There was no significant association between genetically-predicted FGF-23 and LVM index (estimated difference in LVM index per 1-SD higher genetically-predicted logFGF-23 was 0.4g/m^2^ (0.0–0.7; Table 3).

### FGF-23 and risk of non-cardiovascular outcomes

In UK Biobank data, there was no significant association between genetically-predicted FGF-23 with risk of any of the non-cardiovascular outcomes. The ORs per 1-SD higher genetically-predicted logFGF-23 were 1.00 for any fracture (0.97-1.04; n=51,166), 1.08 for any fragility fracture (0.99-1.19; n=6,624), 1.02 for hospitalisation for infection (0.97-1.06; n=38,613), 0.96 for hospitalisation with acute kidney injury (0.89–1.03; n=11,569), 1.02 for treated end-stage kidney disease (0.77-1.34; n=774), and 1.01 for any non-cardiovascular death (0.95-1.07; n=17,196 outcomes; Figure 2).

**Figure 2:**
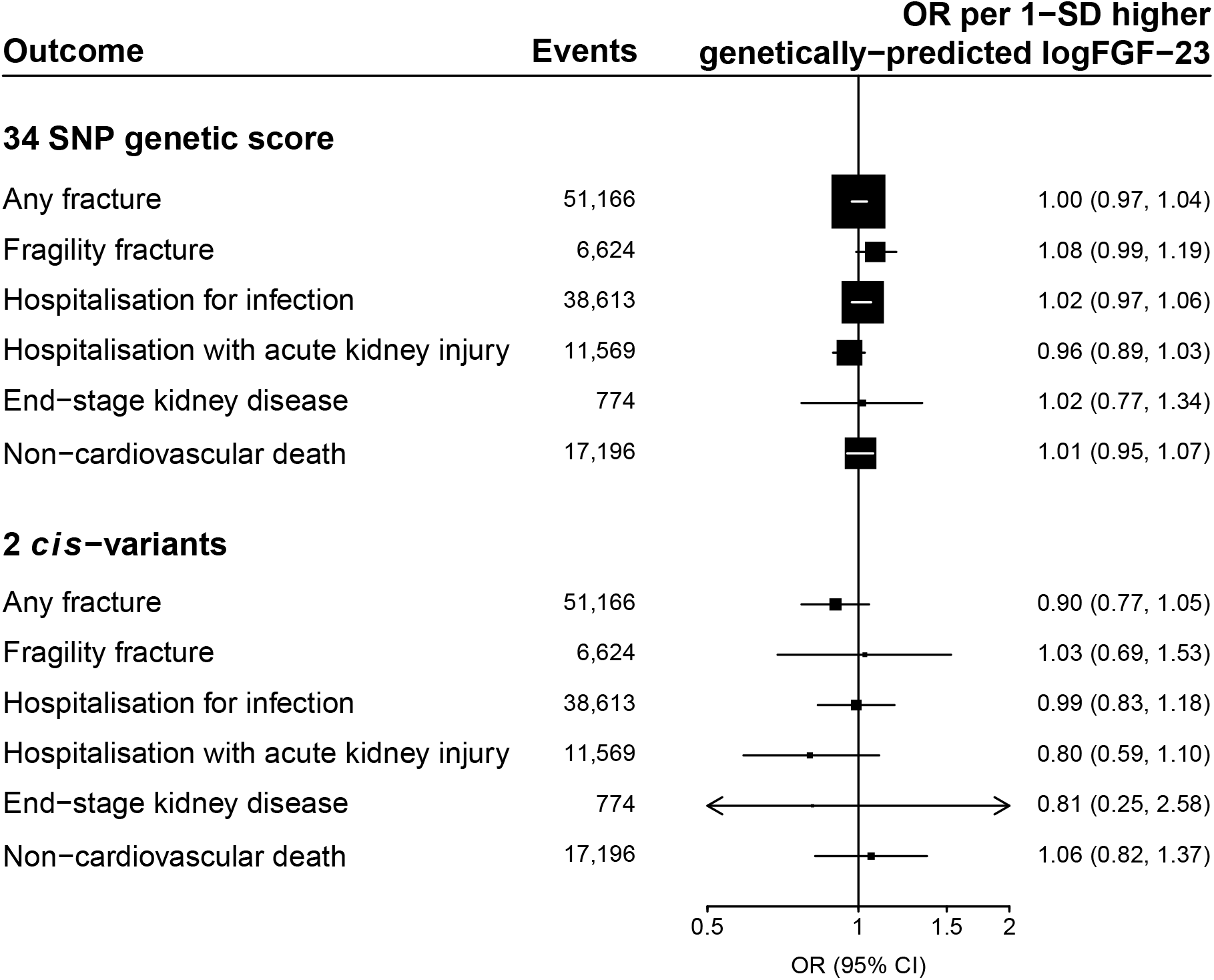
Associations between genetically predicted FGF-23 with risk of non-cardiovascular outcomes. FGF-23, Fibroblast growth factor 23; SNP, single nucleotide polymorphism; OR, odds ratio; SD, standard deviation; 95% CI, 95% confidence interval.

Higher genetically-predicted FGF-23 concentrations were associated with higher gynoid (pelvic girdle) bone mass but not android (lumbar) bone mass, nor bone mineral density in the lumbar vertebrae or femoral neck (Table 3). These associations were no longer significant in a sensitivity analysis excluding 4 SNPs at the *CYP24A1* locus (rs13038432, rs2870308, rs290403, rs1570669), as their associations with FGF-23 may be indirectly mediated via effects on vitamin D metabolism.These associations were also not apparent when using the *cis*-variants alone.

### Sensitivity analyses

Analyses using two *cis-*variants had limited power in UK Biobank (minimum detectable OR with 80% power and α=0.05 for any atherosclerotic event=1.47 per 1-SD higher genetically predicted logFGF-23). Inverse variance weighted MR using these variants yielded an estimated OR per 1-SD higher logFGF-23 of 1.04 (0.84-1.29) for any atherosclerotic and 0.87 (0.64-1.17) for any non-atherosclerotic cardiovascular outcome (Figure 1). Two-sample MR analyses using the case-control consortia provided more power. The minimum detectable ORs for coronary artery disease, stroke and heart failure per FGF-23 increasing allele were 1.13, 1.26 and 1.18, respectively. In such analyses there was no association of genetically predicted FGF-23 with increased risk of coronary artery disease (1.06, 0.95-1.18), ischaemic stroke (0.82, 0.65-1.03) or heart failure (0.82, 0.68-0.98; Figure 1).

Findings were unchanged in a series of other sensitivity analyses using different MR methods (Supplemental Figures 1-5), after exclusion of the 3 SNPs with potential pleiotropy with other cardiovascular traits/risk factors (Supplemental Table 6), after using a more restrictive threshold to define linkage disequilibrium (Supplemental Table 7) and when using a genetic score limited to the 8 variants associated with FGF-23 at p<5×10^−8^ (Supplemental Table 8).

## Discussion

Our overriding aim was to conduct powerful and unconfounded genetic analyses capable of detecting the size of associations between FGF-23 and cardiovascular diseases which have been reported in conventional epidemiological studies.^15^ This was achieved by deriving a novel FGF-23 genetic score from ∼19,000 individuals who have contributed to a large collaborative consortium of studies with genomic and proteomic data, and then performing genetic analyses in large genotyped datasets, including UK Biobank and case-control consortia which have been studying the genetics of cardiovascular disease and provided data on ∼180,000 coronary disease cases, ∼35,000 strokes, and ∼50,000 people with heart failure. Our novel 34-variant FGF-23 genetic score was validated and found to be unconfounded with respect to key confounders, including kidney function. A range of subsequent genetic analyses found no significant associations between genetically-predicted FGF-23 with risk of coronary artery disease, ischaemic stroke or heart failure. Nor were there any significant associations with clinical measurements of carotid artery atherosclerosis or imaging evidence of structural heart disease, indicating that the FGF-23 molecule is unlikely to have a direct causal role in the pathogenesis of cardiovascular disease.

Observational studies in general populations have found an increased risk of myocardial infarction, stroke and heart failure in the order of 40% per 20pg/mL higher FGF-23 concentration (about 1-SD in general populations).^15,45,46^ Used within the available outcome datasets in this report, our FGF-23 genetic instrument was estimated to have 80% power to detect a true increase in the odds of cardiovascular diseases in the order of 5-15%. The lack of any significant genetic FGF-23-cardiovascular disease associations in the presented analyses suggest that, if a causal relationship between FGF-23 and any cardiovascular disease does exist, its size is likely to be substantially smaller than that observed in conventional epidemiological studies. We also did not find any human evidence to support the findings from animal studies that FGF-23 stimulates left ventricular hypertrophy.^47^

Our analyses also had reasonable power to test hypotheses about FGF-23 and risk of certain non-cardiovascular diseases. Contrary to findings from conventional epidemiological studies, we found no evidence for associations between genetically-predicted FGF-23 and risk of infection,^16^ fractures,^17^ or acute kidney injury.^18^ These results further support (albeit indirectly) the theory that residual confounding explains many of the reported positive associations between FGF-23 with risk of many cardiovascular and non-cardiovascular diseases.

In conventional observational studies, associations between FGF-23 and cardiovascular disease are present even when analyses are adjusted for known cardiovascular risk factors and markers of CKD-MBD (PTH, phosphate, calcium).^15^ It is likely therefore that at least some of the residual confounders are unmeasured or unknown variables. There may be unmeasured processes related to calcium/phosphate metabolism which cause cardiovascular disease, and are correlated with FGF-23 levels. FGF-23 in this case may be a marker of these underlying processes, but not a direct cause of cardiovascular disease itself.

Conventional observational studies investigating associations of FGF-23 concentrations and fracture risk and bone mineral density have had conflicting findings.^48,49^ The syndrome of tumour-induced osteomalacia caused by excess FGF-23 production by cancer cells is characterised by impaired bone mineralisation and a propensity to fracture.^50^ Conversely, we observed that higher genetically-predicted FGF-23 was associated with higher gynoid bone mass. These effects were small and inconsistent across different MR methods, but are consistent with findings from another observational study.^48^ A potential mechanism for these conflicting associations is the incompletely understood inter-relationship between vitamin D and FGF-23. Individuals with higher blood vitamin D levels have a tendency towards greater phosphate absorption from the gut, and vitamin D directly induces FGF-23 expression in osteocytes – both mechanisms potentially explaining some observed associations between FGF-23 and higher bone mineral density.^51^ An alternative potential explanation is that individuals with higher bone mass have larger absolute numbers of osteocytes, and greater capacity for FGF-23 production.

The present study benefits from the large scale of data from the SCALLOP Consortium,^28^ UK Biobank,^30^ and the case-control consortia, as well as the ability to better control for residual confounding and reverse causality with genetic epidemiological approaches.^20-22^ Distinct from previous Mendelian randomisation studies of FGF-23 we have sought to distinguish between atherosclerotic and non-atherosclerotic disease and have derived and validated a genetic instrument with greater power than that used previously.^24-27^ However, some limitations exist. First, it was not possible to assess directly the actual difference in FGF-23 in UK Biobank predicted by the genetic risk scores. Instead we provide evidence of the genetic risk score’s validity using data from an independent population where it was shown to predict higher levels of FGF-23. Second, only two *cis*-variants were identified, limiting power for sensitivity analyses using these important, likely more-specific, variants.^39^ Nevertheless, point estimates from analyses using these *cis-*variants were consistent with the results from using the full genetic score (Figure 1). Third, the study was restricted to adults of European ancestry and a general population, meaning results may not be generalisable to other populations. In particular, levels of FGF-23 in patients on maintenance haemodialysis are often 2 orders of magnitude higher than those in general populations.^15^ It is possible disease associations may differ at extremely high concentrations – for example if there is an unusual “threshold” effect above which FGF-23 becomes toxic.

The divergence between the associations of FGF-23 with outcomes observed from conventional epidemiological studies versus using genetic approaches highlights a key challenge to epidemiologists performing observational studies, where it is important to adjust for any degree of kidney disease. Adjusting for kidney function using eGFR is unlikely to fully account for any confounding effect of kidney disease due to a combination of inaccuracy in estimation of kidney function and within-person variability,^52,53^ under-adjustment for the effect of CKD duration, and imprecise adjustment for any effects of kidney disease/dysfunction not captured by eGFR.

In summary, our previous systematic review and meta-analysis of conventional observational studies suggested a lack of exposure-response relationship between FGF-23 and risk of a range of diseases, and that FGF-23 associations are non-specific.^15^ We now demonstrate that genetically-predicted FGF-23 is not associated with risk of atherosclerotic or non-atherosclerotic cardiovascular diseases. The totality of the evidence suggests that FGF-23 does not have a direct causal role in the development of cardiovascular disease, and directly targeting FGF-23 is therefore unlikely to represent a clinically meaningful modifiable target to prevent cardiovascular disease. Conventionally observed associations between FGF-23 with cardiovascular disease are likely due to residual confounding, perhaps due to inability to adjust for kidney disease and any aspects of CKD-mineral bone disease which cause cardiovascular disease.

## Supporting information

STROBE-MR Checklist

## Data Availability

All data produced in the present study are available upon reasonable request to the authors

## Acknowledgements

This research has been conducted using the open-access UK Biobank Resource (application number 15595), with data available from: https://www.ukbiobank.ac.uk/ and in partnership with the SCALLOP consortium (membership listed below). The Medical Research Council Population Health Research Unit at the University of Oxford, which is part of the Clinical Trial Service Unit and Epidemiological Studies Unit (CTSU), receives core funding from the United Kingdom Medical Research Council. Analyses were also supported by Health Data Research UK. KD is funded by an NIHR Academic Clinical Fellowship. WGH is supported by a Medical Research Council Kidney Research UK Professor David Kerr Clinician Scientist Award. MVH works in a unit that receives funding from the UK Medical Research Council and is supported by a British Heart Foundation Intermediate Clinical Research Fellowship (FS/18/23/33512) and the National Institute for Health Research Oxford Biomedical Research Centre. Members of the CVD-I project in the SCALLOP Consortium include: Lasse Folkersen, Stefan Gustafsson, Qin Wang, Jingyuan Fu, Gustav Smith, Tonu Esko, Caroline Hayward, Ulf Gyllensten, Mikael Landen, Agneta Siegbahn, Jim F Wilson, Lars Wallentin, Adam S Butterworth, Michael Holmes, Erik Ingelsson, and Anders Mälarstig

