## Supplemental tables and figures for "Fibroblast Growth Factor-23 and Risk of Cardiovascular Diseases: a Mendelian Randomisation study"

### Supplemental Materials

Supplemental methods: UK Biobank outcome definitions

Supplemental table 1: 34 independent genetic variants for FGF-23 identified in SCALLOP

Supplemental table 2: FGF-23 variants associated with other cardiovascular risk factors (i.e. potentially pleiotropic SNPs)

Supplemental table 3: *Cis*-variant associations with logFGF-23 concentration

Supplemental table 4: Variance explained and minimum detectable ORs for the 34 SNP FGF-23 genetic score in UK Biobank, by outcome

Supplemental table 5: Validation of 34 SNP genetic score using ORIGIN data

Supplemental table 6: Associations between genetically-predicted FGF-23 with risk of outcomes after excluding three SNPs which are potentially pleiotropic with cardiovascular traits/risk factor

Supplemental table 7: Associations between genetically-predicted FGF-23 with risk of outcomes using the 30 FGF-23 SNPs with linkage disequilibrium $r^2<0.001$

Supplemental table 8: Associations between genetically-predicted FGF-23 with risk of outcomes using the 8 FGF-23 SNPs associated with FGF-23 levels at $p<5x10^{-8}$

Supplemental figure 1: Associations between genetically-predicted FGF-23 with risk of atherosclerotic cardiovascular outcomes using standard methods to assess validity of instrumental variable assumptions

Supplemental figure 2: Associations between genetically-predicted FGF-23 with risk of non-atherosclerotic cardiovascular outcomes using standard methods to assess validity of instrumental variable assumptions

Supplemental figure 3: Associations between genetically-predicted FGF-23 with risk of non-cardiovascular outcomes using standard methods to assess validity of instrumental variable assumptions

Supplemental figure 4: Associations between genetically-predicted FGF-23 with clinical measurements using standard methods to assess validity of instrumental variable assumptions

Supplemental figure 5: Scatter plots of effect estimates for individual SNP-FGF-23 associations and associations with key clinical outcomes

Supplemental appendix 1: OPCS-4 codes used to define the outcome “other revascularisation”

Supplemental appendix 2: ICD-10 diagnostic codes used to define hospitalisation for infection

### Supplemental methods: UK Biobank outcome definitions

The following outcomes were defined in UK Biobank using hospital admission and mortality data.

| Outcome | Definition | Notes |
|---|---|---|
| **Any atherosclerotic cardiovascular outcome** | | |
| Non-fatal myocardial infarction | Participant survived 30 days beyond the first day of a hospital episode with an ICD-10 code of I219, I22, I220, I221, I228, I229, I23, I230, I231, I232, I233, I234, I235, I236 or I238 in any diagnostic position | |
| Coronary revascularisation | Participant had a hospital episode with an OPCS-4 code of K49, K50, K75, K40, K41, K42, K43, K44, K45 or K46 | Includes coronary bypass, balloon angioplasty and stenting. |
| Ischaemic stroke (fatal or not) | Participant had a hospital episode with an ICD-10 code of I63 or I64 in any diagnostic position, or died with ICD-10 code I63 or I64 as the underlying cause of death | Includes strokes not specified as haemorrhagic or ischaemic |
| Coronary death | Participant died with an underlying cause of death ICD-10 code of I20, I21, I22, I23, I24, I250, I251, I252, I256, I258 or I259 | Includes ischaemic cardiomyopathy as a cause of death |
| Other revascularisation | Participant had a hospital episode with any of the OPCS-4 procedural codes code listed in Supplemental appendix 1. | Included carotid revascularisation, abdominal viscera/renal revascularisation, upper and lower limb (distal to the aortic bifurcation, including aorto-femoral bypass) revascularisations, but excluded other aortic procedures and amputations |
| **Any non-atherosclerotic cardiovascular outcome** | | |
| Hospitalisation with heart failure | Participant had a hospital episode with an ICD-10 code of I50, I501, I509, I110, I130, I132, I255, I420, I423, I424, I425, I426, I427, I428, I429, I43, I430, I431, I432, or I438 in any diagnostic position | Includes cardiomyopathies due to nutritional, metabolic, infectious and drug causes |
| Non-coronary cardiac death | Participant has died, with the underlying cause of death ICD-10 code of I01, I020, I05, I06, I11, II3, I253, I254, I255, I3, I4, I5 or R96 | Includes unexplained sudden death |
| Other vascular death | Participant has died, with an underlying cause of death ICD-10 code in the range I00-I99, but excluding those already defined as a coronary (see above), other cardiac (see above), or stroke death (I60, I61, I63, I64, I69 and I629) | |
| Haemorrhagic stroke (fatal or not) | Participant had a hospital episode with an ICD-10 code of I60 or I61 in any diagnostic position, or died with an underlying cause of death ICD-10 code of I60 or I61 | Includes subarachnoid haemorrhage |
| **Non-cardiovascular outcomes** | | |
| Any fracture | Participant answered in the affirmative to "Have you fractured a bone in the last 5 years?" at recruitment, or had a hospital episode with a fracture of any bone. | Includes long bones, axial skeleton, hands, feet and skull |
| Fragility fracture | Participant had a hospital episode with an ICD-10 code of M800, M801, M802, M803, M804, M805, M808, M809, M844, S220, S221, S320, S321, S322, S323, S324, S325, S327, S720, S721, or S722 | Includes specified osteoporotic fragility fractures (M80X), fractures of thoracolumbar vertebrae, the pelvic rim and the neck of femur. |

| Outcome | Definition | Notes |
|---|---|---|
| Hospitalisation with acute kidney injury | Participant had a hospital episode with ICD-10 code N17 in any diagnostic position | |
| Hospitalisation for infection | Participant had a hospital episode with an ICD-10 code as listed in Supplemental appendix 2 | Defined as bacteria or parasitic infection that would usually require antimicrobial treatment. Definitions adapted from https://doi.org/10.1136/bmj.k5092 |
| End-stage kidney disease (treated) | Maintenance dialysis or kidney transplant recipient | |
| Non cardiovascular death | Participant has died, with the underlying cause of death ICD-10 which is neither in the range I00-I99, nor R96 | |

**Supplemental table 1: 34 independent genetic variants for FGF-23 identified in SCALLOP**

| SNP rsID | Marker location | Alleles a1 | Alleles a2 | Nearest gene | Minor allele frequency | Association with logFGF-23: Effect size (beta) | Standard error of beta | p value | Number of participants with data |
|---|---|---|---|---|---|---|---|---|---|
| rs6706281 | 2:190452748 | A | C | *SLC40A1* | 0.29 | -0.1057 | 0.0121 | 2.43E-18 | 18239 |
| rs2870308 | 20:52727953 | A | C | *CYP24A1* | 0.27 | -0.0977 | 0.0125 | 4.70E-15 | 18239 |
| rs687289 | 9:136137106 | A | G | *ABO* | 0.37 | 0.0776 | 0.0118 | 5.45E-11 | 15273 |
| rs11748297 | 5:176800361 | A | G | *LMAN2* | 0.29 | -0.0709 | 0.0113 | 3.57E-10 | 19195 |
| *rs6489536* **§** | *12:4491909* | *C* | *G* | *FGF23* | *0.33* | *0.0702* | *0.0122* | *7.77E-09* | *15802* |
| rs4744712 | 9:71434707 | A | C | *FAM122A* | 0.39 | -0.0596 | 0.0104 | 1.08E-08 | 19195 |
| rs34551523 | 5:146629145 | A | G | *STK32A* | 0.03 | 0.2015 | 0.0358 | 1.82E-08 | 15801 |
| rs6561643 | 13:33509079 | A | T | *KL* | 0.37 | -0.0651 | 0.0118 | 3.18E-08 | 18239 |
| rs16988687 | 20:42704488 | T | C | *TOX2* | 0.15 | -0.0785 | 0.0146 | 7.44E-08 | 19195 |
| rs9372822 | 6:125928290 | T | C | *RP11-624M8.1* | 0.35 | -0.0591 | 0.0112 | 1.17E-07 | 19195 |
| rs74461633 | 6:166587846 | T | C | *SNORD45* | 0.04 | 0.1663 | 0.0317 | 1.54E-07 | 15801 |
| rs13038432 | 20:52787302 | A | G | *CYP24A1* | 0.08 | 0.1107 | 0.0214 | 2.30E-07 | 18239 |
| *rs7955866* **§** | *12:4479549* | *A* | *G* | *FGF23* | *0.12* | *-0.0915* | *0.0178* | *2.65E-07* | *15801* |
| rs150865155 | 12:117088921 | A | G | *C12orf49* | 0.01 | -0.3124 | 0.0626 | 5.90E-07 | 13990 |
| rs35827013 | 2:155016134 | A | G | *GALNT13* | 0.27 | 0.0619 | 0.0124 | 6.33E-07 | 19195 |
| rs192539378 | 2:190581687 | A | G | *ANKAR* | 0.05 | -0.1394 | 0.028 | 6.59E-07 | 13932 |
| rs1570669 | 20:52774427 | A | G | *CYP24A1* | 0.34 | -0.0548 | 0.0112 | 9.19E-07 | 18239 |
| rs186557449 | 3:20739233 | T | C | | 0.02 | -0.2606 | 0.0531 | 9.20E-07 | 14501 |
| rs67833371 | 18:8702913 | A | T | *RP11-674N23.1* | 0.37 | 0.0647 | 0.0132 | 1.01E-06 | 12976 |
| rs147720247 | 16:65073369 | T | G | *CDH11* | 0.04 | 0.1493 | 0.0311 | 1.59E-06 | 17326 |
| rs11035939 | 11:40600846 | A | G | *LRRC4C* | 0.28 | -0.0599 | 0.0125 | 1.77E-06 | 15801 |
| rs189972262 | 4:12448579 | T | C | | 0.02 | 0.3595 | 0.0754 | 1.86E-06 | 9788 |
| rs2720020 | 3:163629696 | T | G | | 0.21 | 0.0651 | 0.0137 | 1.86E-06 | 19195 |
| rs147321547 | 19:51883001 | T | C | *LIM2* | 0.19 | -0.0705 | 0.0148 | 2.03E-06 | 15801 |
| rs117612483 | 13:111392745 | T | G | *ING1* | 0.02 | 0.2503 | 0.0533 | 2.63E-06 | 14624 |
| rs290403 | 20:52711514 | A | G | *CYP24A1* | 0.47 | 0.0524 | 0.0112 | 2.93E-06 | 18239 |
| rs78450448 | 15:53626754 | T | C | *WDR72* | 0.02 | 0.202 | 0.0432 | 2.96E-06 | 15457 |
| rs117989952 | 8:4020568 | A | G | *CSMD1* | 0.01 | 0.5645 | 0.1209 | 3.05E-06 | 8067 |

| SNP rsID | Marker location | Alleles | | Nearest gene | Minor allele frequency | Association with logFGF-23 | | | Number of participants with data |
|---|---|---|---|---|---|---|---|---|---|
| | | a1 | a2 | | | Effect size (beta) | Standard error of beta | p value | |
| rs75357988 | 5:161568285 | T | C | *GABRG2* | 0.02 | -0.225 | 0.0483 | 3.19E-06 | 14390 |
| rs11542063 | 15:45353365 | A | G | *SORD* | 0.02 | 0.2488 | 0.0536 | 3.45E-06 | 14624 |
| rs61855139 | 10:70223133 | A | G | *DNA2* | 0.10 | -0.0888 | 0.0193 | 4.33E-06 | 17326 |
| rs79146532 | 17:34934676 | A | G | *GGNBP2* | 0.03 | -0.1618 | 0.0353 | 4.41E-06 | 19195 |
| rs9695235 | 9:119138128 | T | C | *PAPPA* | 0.20 | 0.0611 | 0.0134 | 4.77E-06 | 19195 |
| rs11217709 | 11:120029788 | T | C | *TRIM29* | 0.04 | -0.1539 | 0.0337 | 4.83E-06 | 14845 |

FGF-23, Fibroblast growth factor 23; SNP, single nucleotide polymorphism. § denotes the 2 *cis*-variants within 100kb of the FGF-23 transcription start site. Marker locations uses GRCh37 co-ordinates. Allele1 is the effect allele. Effects are measured in standard deviations of $log_2$ transformed FGF-23.

**Supplemental table 2: FGF-23 variants associated with other cardiovascular trait/risk factors (i.e. potentially pleiotropic SNPs)**

| SNP rsID | Alleles a1 | Alleles a2 | Trait/ risk factor | Data source | Ancestry | Year | Association with trait/risk factor: Effect size (beta) | Standard error of beta | p value | Direction | no. with data | no. of studies | unit of analysis |
|---|---|---|---|---|---|---|---|---|---|---|---|---|---|
| rs687289 | G | A | Low density lipoprotein | PMID 24097068 | European | 2013 | -0.0403 | 0.004 | 1.33E-24 | - | 172684 | 60 | IVNT |
| | G | A | Low density lipoprotein | PMID 20686565 | Mixed | 2010 | -0.0433 | 0.005 | 5.74E-16 | - | 95454 | 46 | z score |
| | G | A | Total cholesterol | PMID 24097068 | European | 2013 | -0.0403 | 0.004 | 2.14E-26 | - | 186901 | 60 | IVNT |
| | G | A | Total cholesterol | PMID 20686565 | Mixed | 2010 | -0.0408 | 0.0052 | 6.09E-15 | - | 100184 | 46 | z score |
| | G | A | Diastolic BP | UK Biobank | European | 2017 | 0.01664 | 0.002643 | 3.09E-10 | + | 317756 | 1 | IVNT |
| | G | A | Doctor diagnosed : hypertension | UK Biobank | European | 2017 | 0.006404 | 0.001161 | 3.49E-08 | + | 336683 | 1 | Risk difference |
| rs6561643 | A | T | Type 2 DM | PMID 28566273 | European | 2017 | 0.04 | 0.014 | 0.004 | + | 159208 | 18 | logOR |
| rs79146532 | A | G | Tobacco smoking: (occasional) | UK Biobank | European | 2017 | -0.00536 | 0.001751 | 0.002 | - | 83133 | 1 | Risk difference |

FGF-23, Fibroblast growth factor 23; SNP, single nucleotide polymorphism; BP, blood pressure; DM, diabetes mellitus; PMID, Pubmed ID; IVNT= inverse normally ranked phenotype; OR=odds ratio. Associations with cardiovascular traits/risk factors for the 34x FGF-23 SNPs (and variants in linkage disequilibrium [$r^2$>0.8; 1000 Genomes Phase 3 release]) identified from published data from PhenoScanner (http://www.phenoscanner.medschl.cam.ac.uk/about/)

**Supplemental table 3: *Cis*-variant associations with logFGF-23 concentration**

| SNP rsID | Increase in logFGF-23 per effect allele (standard error) | p values | |
|---|---|---|---|
| | | univariate | conditioned on other *cis*-variant |
| rs6489536 | 0.0702 (0.012) | $7.8x10^{-9}$ | $5.4x10^{-6}$ |
| rs7955866 | -0.0915 (0.0178) | $2.7x10^{-7}$ | $1.8x10^{-4}$ |

FGF-23, Fibroblast growth factor 23; SNP, single nucleotide polymorphism

**Supplemental table 4: Variance explained and minimum detectable ORs for the 34 SNP FGF-23 genetic score in UK Biobank, by outcome**

| | Proportion of participants with outcome | Proportion of variance of logFGF-23 explained | Minimal detectable OR per 1-SD higher logFGF-23 with 80% power, 2p=0.05 |
|---|---|---|---|
| **Atherosclerotic cardiovascular event** | | | |
| Non-fatal myocardial infarction | 0.029 | | 1.12 |
| Ischaemic stroke | 0.018 | | 1.15 |
| Coronary revascularisation | 0.043 | 0.063 | 1.10 |
| Other revascularisation | 0.011 | | 1.19 |
| Coronary death | 0.007 | | 1.24 |
| **Any atherosclerotic cardiovascular event** | **0.078** | | **1.08** |
| **Non-atherosclerotic cardiovascular outcomes** | | | |
| Hospitalisation with heart failure | 0.030 | | 1.12 |
| Haemorrhagic stroke | 0.005 | | 1.28 |
| Non-coronary cardiac death | 0.002 | 0.063 | 1.44 |
| Other vascular death | 0.002 | | 1.44 |
| **Any non-atherosclerotic cardiovascular event** | **0.037** | | **1.11** |
| **Non-cardiovascular outcomes** | | | |
| Any fracture | 0.15 | | 1.06 |
| Fragility fracture | 0.020 | | 1.14 |
| Hospitalisation for infection | 0.114 | | 1.07 |
| Hospitalisation with acute kidney injury | 0.034 | 0.063 | 1.12 |
| End-stage kidney disease (treated) | 0.002 | | 1.36 |
| Non-cardiovascular death | 0.054 | | 1.09 |

FGF-23, Fibroblast growth factor 23; OR, odds ratio; SNP, single nucleotide polymorphism; SD, standard deviation

**Supplemental table 5: Validation of 34 SNP genetic score using ORIGIN data**

| Range of FGF23 concentrations (pg/mL) | No. of participants | OR (95% CI) of being in this category or a higher category, per 1 unit higher genetic score | p value |
|---|---|---|---|
| <49 | 2575 | -- | |
| [49 – 60] | 430 | 1.05 (0.99 – 1.12) | |
| [60 – 80] | 485 | 1.07 (1.00 – 1.15) | |
| [80 – 120] | 389 | 1.10 (1.02 – 1.18) | |
| [120 – 4500] | 511 | 1.13 (1.03 – 1.24) | |
| **OVERALL** | **4390** | **1.07 (1.00 – 1.13)** | **0.036** |

FGF-23, Fibroblast growth factor 23; SNP= single nucleotide polymorphism; OR (95% CI), odds ratio (95% confidence interval). Analyses use ordinal regression adjusted for age, sex and ethnicity. Ordinal regression was used due to the high proportion of participants with values of FGF-23 below the lower limit of detection for the assay.

**Supplemental table 6: Associations between genetically-predicted FGF-23 with risk of outcomes after excluding three SNPs which are potentially pleiotropic with cardiovascular traits/risk factor**

| Outcome | Number of outcomes | OR per 1-SD higher genetically-predicted logFGF-23 (95% CI) | p-value (Bonferroni corrected) |
|---|---|---|---|
| **Atherosclerotic cardiovascular outcomes** | | | |
| Non-fatal myocardial infarction | 9,677 | 1.05 (0.96 – 1.14) | 0.99 |
| Ischaemic stroke | 5,992 | 1.01 (0.91 – 1.12) | 0.99 |
| Coronary revascularisation | 14,646 | 1.01 (0.95 – 1.09) | 0.98 |
| Other revascularisation | 3,782 | 0.96 (0.84 – 1.09) | 0.99 |
| Coronary death | 2,258 | 1.13 (0.95 – 1.34) | 0.99 |
| **Any atherosclerotic cardiovascular event** | **26,266** | **1.01 (0.96 – 1.07)** | **0.99** |
| **Non-atherosclerotic cardiovascular outcomes** | | | |
| Hospitalisation with heart failure | 10,177 | 1.00 (0.92 – 1.09) | 0.99 |
| Haemorrhagic stroke | 1,745 | 1.06 (0.87 – 1.28) | 0.99 |
| Non-coronary cardiac death | 689 | 0.96 (0.70 – 1.30) | 0.99 |
| Other vascular death | 617 | 0.95 (0.68 – 1.31) | 0.99 |
| **Any non-atherosclerotic cardiovascular event** | **12,652** | **1.01 (0.93 – 1.08)** | **0.99** |
| **Non-cardiovascular outcomes** | | | |
| Any fracture | 51,166 | 1.00 (0.96 – 1.04) | 0.99 |
| Fragility fracture | 6,624 | 1.05 (0.95 – 1.16) | 0.99 |
| Hospitalisation for infection | 38,613 | 1.01 (0.97 – 1.06) | 0.99 |
| Hospitalisation with acute kidney injury | 11,569 | 0.96 (0.89 – 1.04) | 0.99 |
| End-stage kidney disease (treated) | 774 | 1.04 (0.77 – 1.38) | 0.99 |
| Non-vascular death | 17,196 | 1.00 (0.94 – 1.07) | 0.99 |
| **Clinical measurement (units)** | **Number with measurement** | **Effect of 1-SD higher genetically predicted logFGF-23 (95% CI)** | |
| Android bone mass (grams) | 3,695 | 1.43 (-0.04,2.91) | 0.73 |
| Gynoid bone mass (grams) | 3,695 | 9.29 (3.43,15.14) | 0.04 |
| Lumbar vertebral bone mineral density (g/cm$^3$) | 3,679 | 0.027 (0.002,0.052) | 0.57 |
| Femoral neck bone mineral density (g/cm$^3$) | 3,704 | 0.018 (0.000,0.036) | 0.71 |
| Carotid intimal thickness (maximum; μm) | 31,641 | -0.3 (-6.4,5.8) | 0.99 |
| Carotid intimal thickness (mean; μm) | 31,641 | -1.5 (-6.6,3.6) | 0.99 |
| Left ventricular mass index (g/m$^2$) | 18,710 | 0.26 (-0.13,0.65) | 0.99 |

FGF-23, Fibroblast growth factor 23; SNP= single nucleotide polymorphism; OR (95% CI), odds ratio (95% confidence interval); SD, standard deviation. Excluded SNP rsID's: rs687289, rs6561643 and rs79146532

**Supplemental table 7: Associations between genetically-predicted FGF-23 with risk of outcomes using the 30 FGF-23 SNPs with linkage disequilibrium $r^2$<0.001**

| Outcome | Number of outcomes | OR per 1-SD higher genetically-predicted logFGF-23 (95% CI) | p-value (Bonferroni corrected) |
|---|---|---|---|
| **Atherosclerotic cardiovascular outcomes** | | | |
| Non-fatal myocardial infarction | 9,677 | 1.07 (0.98 – 1.17) | 0.92 |
| Ischaemic stroke | 5,992 | 1.03 (0.92 – 1.15) | 0.99 |
| Coronary revascularisation | 14,646 | 1.02 (0.94 – 1.09) | 0.99 |
| Other revascularisation | 3,782 | 0.95 (0.83 – 1.10) | 0.99 |
| Coronary death | 2,258 | 1.15 (0.96 – 1.38) | 0.94 |
| **Any atherosclerotic cardiovascular event** | **26,266** | **1.02 (0.97 – 1.08)** | **0.99** |
| **Non-atherosclerotic cardiovascular outcomes** | | | |
| Hospitalisation with heart failure | 10,177 | 1.01 (0.93 – 1.10) | 0.99 |
| Haemorrhagic stroke | 1,745 | 1.04 (0.85 – 1.28) | 0.99 |
| Non-coronary cardiac death | 689 | 0.95 (0.69 – 1.31) | 0.99 |
| Other vascular death | 617 | 0.94 (0.67 – 1.33) | 0.99 |
| **Any non-atherosclerotic cardiovascular event** | **12,652** | **1.01 (0.93 – 1.09)** | **0.99** |
| **Non-cardiovascular outcomes** | | | |
| Any fracture | 51,166 | 1.01 (0.97 – 1.05) | 0.99 |
| Fragility fracture | 6,624 | 1.08 (0.98 – 1.21) | 0.96 |
| Hospitalisation for infection | 38,613 | 1.02 (0.97 – 1.07) | 0.99 |
| Hospitalisation with acute kidney injury | 11,569 | 0.95 (0.88 – 1.03) | 0.99 |
| End-stage kidney disease (treated) | 774 | 1.00 (0.73 – 1.35) | 0.99 |
| Non-cardiovascular death | 17,196 | 1.00 (0.93 – 1.07) | 0.99 |
| **Clinical measurement (units)** | **Number with measurement** | **Effect of 1-SD higher genetically predicted logFGF-23 (95% CI)** | |
| Android bone mass (g) | 3,695 | 1.21 (-0.34,2.75) | 0.95 |
| Gynoid bone mass (g) | 3,695 | 8.28 (2.14,14.42) | 0.17 |
| Lumbar vertebral bone mineral density (g/cm$^3$) | 3,679 | 0.026 (-0.001,0.053) | 0.72 |
| Femoral neck bone mineral density (g/cm$^3$) | 3,704 | 0.018 (-0.001,0.037) | 0.76 |
| Carotid intimal thickness (maximum; $\mu$m) | 31,641 | -0.3 (-6.8,6.1) | 0.99 |
| Carotid intimal thickness (mean; $\mu$m) | 31,641 | -1.2 (-6.5,4.2) | 0.99 |
| Left ventricular mass index (g/m$^2$) | 18,710 | 0.46 (0.05,0.87) | 0.44 |

FGF-23, Fibroblast growth factor 23; SNP= single nucleotide polymorphism; OR (95% CI), odds ratio (95% confidence interval); SD, standard deviation.

**Supplemental table 8: Associations between genetically-predicted FGF-23 with risk of outcomes using the 8 FGF-23 SNPs associated with FGF-23 levels at $p<5x10^{-8}$**

| Outcome | Number of outcomes | OR per 1-SD higher genetically-predicted logFGF-23 (95% CI) | p-value (Bonferroni corrected) |
|---|---|---|---|
| **Atherosclerotic cardiovascular outcomes** | | | |
| Non-fatal myocardial infarction | 9,677 | 1.15 (1.00 – 1.32) | 0.72 |
| Ischaemic stroke | 5,992 | 0.98 (0.82 – 1.18) | 0.99 |
| Coronary revascularisation | 14,646 | 1.07 (0.95 – 1.20) | 0.99 |
| Other revascularisation | 3,782 | 1.04 (0.83 – 1.30) | 0.73 |
| Coronary death | 2,258 | 1.08 (0.81 – 1.44) | 0.99 |
| **Any atherosclerotic cardiovascular event** | **26,266** | **1.06 (0.97 – 1.16)** | **0.99** |
| **Non-atherosclerotic cardiovascular outcomes** | | | |
| Hospitalisation with heart failure | 10,177 | 1.01 (0.88 – 1.16) | 0.99 |
| Haemorrhagic stroke | 1,745 | 1.00 (0.72 – 1.39) | 0.99 |
| Non-coronary cardiac death | 689 | 0.68 (0.40 – 1.15) | 0.97 |
| Other vascular death | 617 | 0.93 (0.54 – 1.62) | 0.99 |
| **Any non-atherosclerotic cardiovascular event** | **12,652** | **0.98 (0.87 – 1.12)** | **0.99** |
| **Non-cardiovascular outcomes** | | | |
| Any fracture | 51,166 | 1.03 (0.97 – 1.10) | 0.99 |
| Fragility fracture | 6,624 | 1.29 (1.09 – 1.53) | 0.07 |
| Hospitalisation for infection | 38,613 | 1.07 (0.99 – 1.15) | 0.84 |
| Hospitalisation with acute kidney injury | 11,569 | 0.97 (0.85 – 1.10) | 0.99 |
| End-stage kidney disease (treated) | 774 | 1.02 (0.62 – 1.67) | 0.99 |
| Non-cardiovascular death | 17,196 | 1.02 (0.92 – 1.14) | 0.99 |
| **Clinical measurement (units)** | **Number with measurement** | **Effect of 1-SD higher genetically predicted logFGF-23 (95% CI)** | |
| Android bone mass (g) | 3,695 | -0.31 (-2.83,2.21) | 0.99 |
| Gynoid bone mass (g) | 3,695 | 4.01 (-5.97,14.04) | 0.99 |
| Lumbar vertebral bone mineral density ($g/cm^3$) | 3,679 | 0.005 (-0.039,0.048) | 0.99 |
| Femoral neck bone mineral density ($g/cm^3$) | 3,704 | 0.005 (-0.026,0.036) | 0.99 |
| Carotid intimal thickness (maximum; μm) | 31,641 | 0.3 (-10.1,10.7) | 0.99 |
| Carotid intimal thickness (mean; μm) | 31,641 | 0.4 (-8.3,9.1) | 0.99 |
| Left ventricular mass index ($g/m^2$) | 18,710 | 0.99 (0.33,1.65) | 0.07 |

FGF-23, Fibroblast growth factor 23. The 8 contributory SNPs are indicated in Supplemental Table 1

**Supplemental figure 1: Associations between genetically−predicted FGF−23 with risk of atherosclerotic cardiovascular outcomes using standard methods to assess validity of instrumental variable assumptions (sensitivity analysis)**

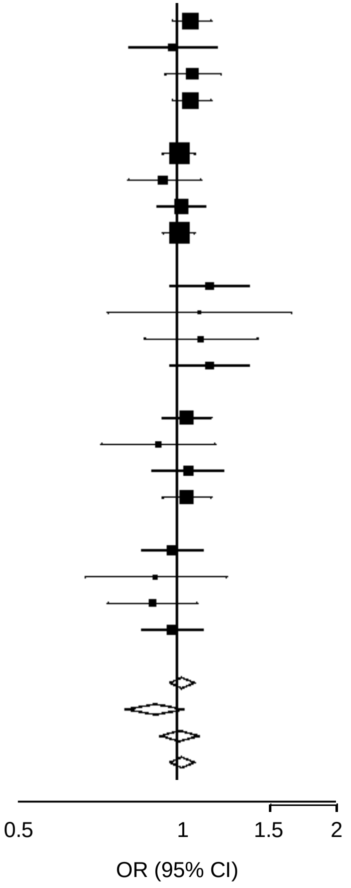


| Outcome | Method | OR (95% CI) per 1−SD increase in genetically−predicted logFGF−23 | p−value | Intercept | SE intercept | p−value intercept |
|---|---|---|---|---|---|---|
| Non−fatal myocardial infarction | IVW | 1.06 (0.98, 1.16) | 0.167 | | | |
| | MR−Egger | 0.98 (0.81, 1.19) | 0.844 | 0.008 | 0.009 | 0.372 |
| | Weighted median | 1.07 (0.95, 1.21) | 0.265 | | | |
| | Outlier corrected IVW | 1.06 (0.98, 1.16) | 0.167 | | | |
| Coronary revascularisation | IVW | 1.01 (0.94, 1.08) | 0.818 | | | |
| | MR−Egger | 0.94 (0.81, 1.11) | 0.489 | 0.006 | 0.007 | 0.379 |
| | Weighted median | 1.02 (0.92, 1.13) | 0.697 | | | |
| | Outlier corrected IVW | 1.01 (0.94, 1.08) | 0.818 | | | |
| Coronary death | IVW | 1.15 (0.97, 1.37) | 0.116 | | | |
| | MR−Egger | 1.10 (0.74, 1.64) | 0.636 | 0.004 | 0.018 | 0.817 |
| | Weighted median | 1.11 (0.87, 1.42) | 0.414 | | | |
| | Outlier corrected IVW | 1.15 (0.97, 1.37) | 0.116 | | | |
| Ischaemic stroke | IVW | 1.04 (0.94, 1.16) | 0.439 | | | |
| | MR−Egger | 0.92 (0.72, 1.18) | 0.521 | 0.012 | 0.011 | 0.276 |
| | Weighted median | 1.05 (0.90, 1.22) | 0.529 | | | |
| | Outlier corrected IVW | 1.04 (0.94, 1.16) | 0.439 | | | |
| Other revascularisation | IVW | 0.98 (0.86, 1.12) | 0.805 | | | |
| | MR−Egger | 0.91 (0.67, 1.24) | 0.554 | 0.007 | 0.014 | 0.590 |
| | Weighted median | 0.90 (0.74, 1.09) | 0.288 | | | |
| | Outlier corrected IVW | 0.98 (0.86, 1.12) | 0.805 | | | |
| **Any atherosclerotic cardiovascular event** | **IVW** | **1.02 (0.97, 1.08)** | **0.397** | | | |
| | **MR−Egger** | **0.91 (0.80, 1.03)** | **0.142** | **0.011** | **0.006** | **0.041** |
| | **Weighted median** | **1.01 (0.93, 1.10)** | **0.776** | | | |
| | **Outlier corrected IVW** | **1.02 (0.97, 1.08)** | **0.397** | | | |

OR (95% CI), odds ratio (95% confidence interval); SD, standard deviation. SE, standard error; IVW=inverse variance weighted. Outlier corrected IVW used the modified Q statistic.

**Supplemental figure 2: Associations between genetically−predicted FGF−23 with risk of non−atherosclerotic cardiovascular outcomes using standard methods to assess validity of instrumental variable assumptions (sensitivity analysis)**

| Outcome | Method | OR (95% CI) per 1−SD increase in genetically−predicted logFGF−23 | p−value | Intercept | SE intercept | p−value intercept |
|---|---|---|---|---|---|---|
| Hospitalisation for heart failure | IVW | 1.01 (0.93, 1.10) | 0.762 | | | |
| | MR−Egger | 1.12 (0.93, 1.35) | 0.242 | −0.010 | 0.008 | 0.249 |
| | Weighted median | 0.97 (0.87, 1.09) | 0.656 | | | |
| | Outlier corrected IVW | 1.01 (0.93, 1.10) | 0.762 | | | |
| Other (non−coronary) cardiac death | IVW | 0.92 (0.68, 1.25) | 0.601 | | | |
| | MR−Egger | 0.82 (0.43, 1.59) | 0.562 | 0.012 | 0.030 | 0.702 |
| | Weighted median | 0.77 (0.49, 1.19) | 0.242 | | | |
| | Outlier corrected IVW | 0.92 (0.68, 1.25) | 0.601 | | | |
| Haemorrhagic stroke | IVW | 1.06 (0.88, 1.30) | 0.528 | | | |
| | MR−Egger | 1.07 (0.68, 1.69) | 0.762 | −0.001 | 0.020 | 0.973 |
| | Weighted median | 0.93 (0.70, 1.23) | 0.599 | | | |
| | Outlier corrected IVW | 1.06 (0.88, 1.30) | 0.528 | | | |
| Other vascular death | IVW | 0.93 (0.67, 1.28) | 0.641 | | | |
| | MR−Egger | 1.80 (0.88, 3.71) | 0.110 | −0.066 | 0.033 | 0.043 |
| | Weighted median | 0.85 (0.54, 1.35) | 0.493 | | | |
| | Outlier corrected IVW | 0.93 (0.67, 1.28) | 0.641 | | | |
| **Any non−atherosclerotic cardiovascular event** | **IVW** | **1.02 (0.94, 1.09)** | **0.685** | | | |
| | **MR−Egger** | **1.13 (0.95, 1.34)** | **0.155** | **−0.011** | **0.008** | **0.166** |
| | **Weighted median** | **0.98 (0.88, 1.09)** | **0.768** | | | |
| | **Outlier corrected IVW** | **1.02 (0.94, 1.09)** | **0.685** | | | |

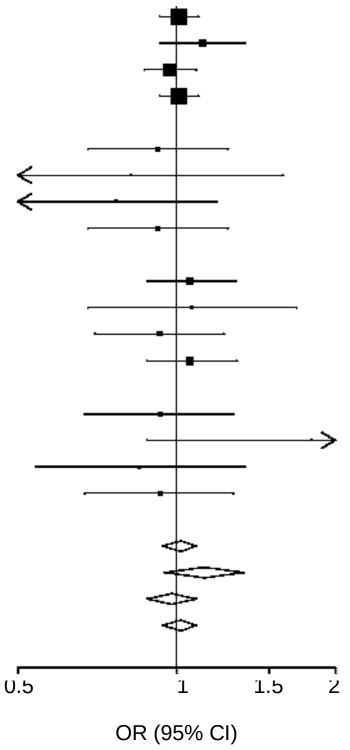


OR (95% CI), odds ratio (95% confidence interval); SD, standard deviation. SE, standard error; IVW=inverse variance weighted. Outlier corrected IVW used the modified Q statistic.

**Supplemental figure 3: Associations between genetically−predicted FGF−23 with risk of non−cardiovascular outcomes using standard methods to assess validity of instrumental variable assumptions (sensitivity analysis)**

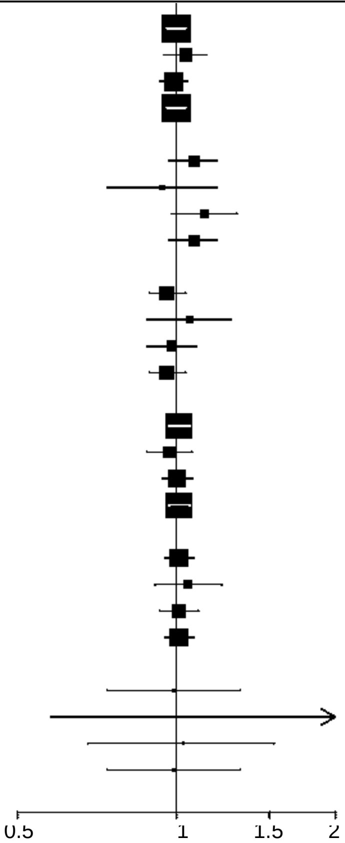


| Outcome | Method | OR (95% CI) per 1−SD increase in genetically−predicted logFGF−23 | p−value | Intercept | SE intercept | p−value intercept |
|---|---|---|---|---|---|---|
| Any fracture | IVW | 1.00 (0.96, 1.04) | 0.928 | | | |
| | MR−Egger | 1.04 (0.95, 1.14) | 0.407 | −0.004 | 0.004 | 0.381 |
| | Weighted median | 0.99 (0.93, 1.05) | 0.733 | | | |
| | Outlier corrected IVW | 1.00 (0.96, 1.04) | 0.928 | | | |
| Fragility fracture | IVW | 1.08 (0.97, 1.19) | 0.162 | | | |
| | MR−Egger | 0.94 (0.74, 1.19) | 0.607 | 0.013 | 0.010 | 0.215 |
| | Weighted median | 1.13 (0.98, 1.30) | 0.104 | | | |
| | Outlier corrected IVW | 1.08 (0.97, 1.19) | 0.162 | | | |
| Hospitalisation with acute kidney injury | IVW | 0.96 (0.89, 1.04) | 0.297 | | | |
| | MR−Egger | 1.06 (0.88, 1.27) | 0.542 | −0.009 | 0.008 | 0.238 |
| | Weighted median | 0.98 (0.88, 1.09) | 0.693 | | | |
| | Outlier corrected IVW | 0.96 (0.89, 1.04) | 0.297 | | | |
| Hospitalisation for infection | IVW | 1.01 (0.97, 1.06) | 0.577 | | | |
| | MR−Egger | 0.97 (0.88, 1.07) | 0.542 | 0.004 | 0.005 | 0.343 |
| | Weighted median | 1.00 (0.94, 1.07) | 0.957 | | | |
| | Outlier corrected IVW | 1.01 (0.97, 1.06) | 0.577 | | | |
| Non−cardiovascular death | IVW | 1.01 (0.95, 1.08) | 0.720 | | | |
| | MR−Egger | 1.05 (0.91, 1.22) | 0.483 | −0.004 | 0.006 | 0.544 |
| | Weighted median | 1.01 (0.93, 1.10) | 0.835 | | | |
| | Outlier corrected IVW | 1.01 (0.95, 1.08) | 0.720 | | | |
| End stage kidney disease (treated) | IVW | 0.99 (0.74, 1.32) | 0.926 | | | |
| | MR−Egger | 1.14 (0.58, 2.21) | 0.704 | −0.014 | 0.030 | 0.640 |
| | Weighted median | 1.03 (0.68, 1.53) | 0.904 | | | |
| | Outlier corrected IVW | 0.99 (0.74, 1.32) | 0.926 | | | |

FGF-23, Fibroblast growth factor 23; OR (95% CI), odds ratio (95% confidence interval); SD, standard deviation. SE, standard error; IVW=inverse variance weighted. Outlier corrected IVW used the modified Q statistic

### Supplemental figure 4: Assocaitions between genetically-predicted FGF-23 with clinical measurements using standard methods to assess validity of instrumental variable assumptions (sensitivity analysis)

**A)**

| Outcome | Method | Effect (95% CI) per 1−SD increase in genetically−predicted logFGF−23 | p−value | Intercept | SE intercept | p−value intercept |
|---|---|---|---|---|---|---|
| **Carotid intimal thickness (μm)** | | | | | | |
| Maximum | IVW | 0.20 (−6.00, 6.40) | 0.940 | | | |
| | MR−Egger | 2.70 (−11.50, 16.90) | 0.708 | −0.241 | 0.635 | 0.704 |
| | Weighted median | −0.30 (−8.90, 8.30) | 0.945 | | | |
| | Outlier corrected IVW | 0.20 (−6.00, 6.40) | 0.940 | | | |
| Mean | IVW | −0.60 (−5.80, 4.60) | 0.810 | | | |
| | MR−Egger | 1.20 (−10.60, 13.10) | 0.839 | −0.182 | 0.531 | 0.732 |
| | Weighted median | −1.20 (−8.60, 6.10) | 0.746 | | | |
| | Outlier corrected IVW | −0.60 (−5.80, 4.60) | 0.810 | | | |

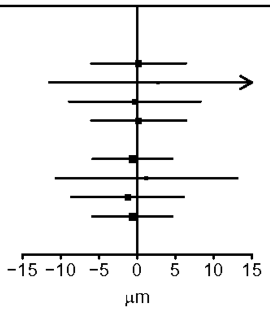


**B)**

| Outcome | Method | Effect (95% CI) per 1−SD increase in genetically−predicted logFGF−23 | p−value | Intercept | SE intercept | p−value intercept |
|---|---|---|---|---|---|---|
| **Left ventricular mass index (g/m$^2$)** | | | | | | |
| | IVW | 0.40 (0.00, 0.80) | 0.068 | | | |
| | MR−Egger | −0.80 (−1.70, 0.20) | 0.106 | 0.109 | 0.041 | 0.007 |
| | Weighted median | 0.40 (−0.20, 1.00) | 0.167 | | | |
| | Outlier corrected IVW | 0.40 (0.00, 0.80) | 0.068 | | | |

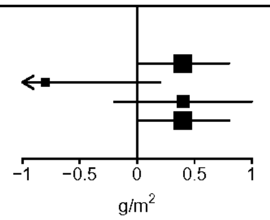


**C)**

| Outcome | Method | Effect (95% CI) per 1−SD increase in genetically−predicted logFGF−23 | p−value | Intercept | SE intercept | p−value intercept |
|---|---|---|---|---|---|---|
| **Bone mass (grams)** | | | | | | |
| Android | IVW | 1.30 (−0.20, 2.80) | 0.091 | | | |
| | MR−Egger | 0.20 (−3.20, 3.70) | 0.892 | 0.103 | 0.154 | 0.506 |
| | Weighted median | 1.20 (−0.90, 3.30) | 0.264 | | | |
| | Outlier corrected IVW | 1.30 (−0.20, 2.80) | 0.091 | | | |
| Gynoid | IVW | 8.60 (2.70, 14.60) | 0.005 | | | |
| | MR−Egger | 4.70 (−9.10, 18.40) | 0.505 | 0.384 | 0.611 | 0.530 |
| | Weighted median | 6.40 (−2.40, 15.20) | 0.154 | | | |
| | Outlier corrected IVW | 8.60 (2.70, 14.60) | 0.005 | | | |

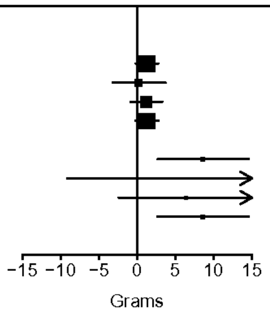


**D)**

| Outcome | Method | Effect (95% CI) per 1−SD increase in genetically−predicted logFGF−23 | p−value | Intercept | SE intercept | p−value intercept |
|---|---|---|---|---|---|---|
| **Bone mineral density (g/cm$^3$)** | | | | | | |
| Lumbar vertebrae | IVW | 0.02 (0.00, 0.05) | 0.063 | | | |
| | MR−Egger | 0.02 (−0.04, 0.08) | 0.496 | 0.000 | 0.003 | 0.889 |
| | Weighted median | 0.02 (−0.02, 0.05) | 0.356 | | | |
| | Outlier corrected IVW | 0.02 (0.00, 0.05) | 0.063 | | | |
| Femoral neck | IVW | 0.02 (0.00, 0.04) | 0.067 | | | |
| | MR−Egger | 0.05 (0.01, 0.09) | 0.028 | −0.003 | 0.002 | 0.116 |
| | Weighted median | 0.01 (−0.02, 0.04) | 0.545 | | | |
| | Outlier corrected IVW | 0.02 (0.00, 0.04) | 0.067 | | | |

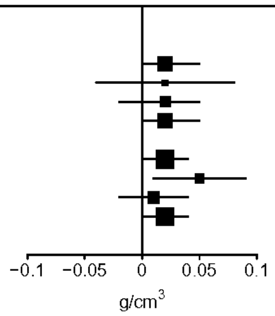


SD, standard deviation. SE, standard error; IVW=inverse variance weighted. Outlier corrected IVW used the modified Q statistic.

**Supplemental figure 5: Scatter plots of effect estimates for individual SNP-FGF-23 associations and associations with key clinical outcomes**

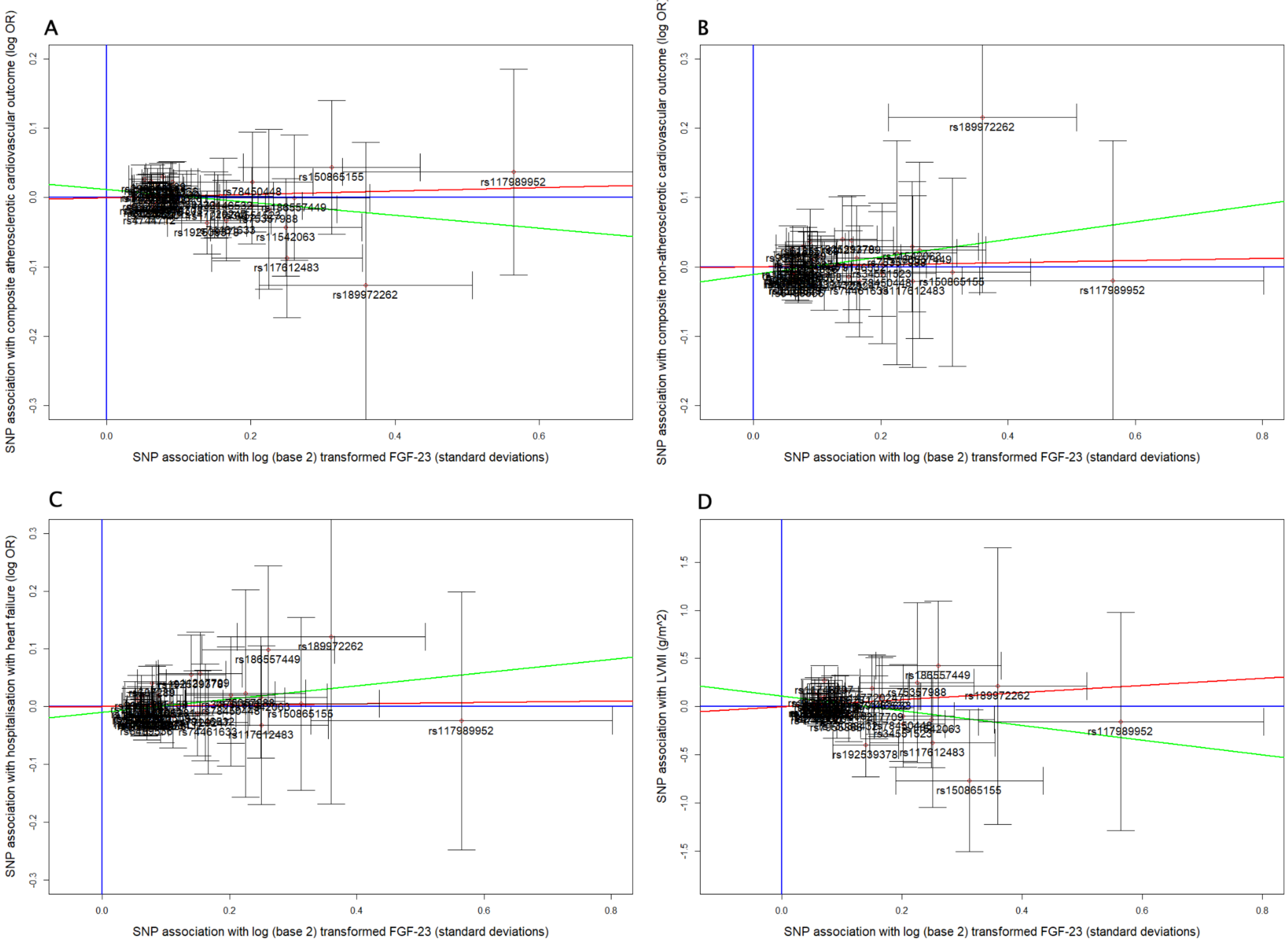


FGF-23, Fibroblast growth factor 23; Individual SNP estimates and 95% CIs for FGF-23 associations and associations with the composite atherosclerotic clinical outcome (A), the composite non-atherosclerotic clinical outcome (B), hospitalisation with heart-failure (C) and LV mass index (D). Inverse-variance weighted regression lines are shown in red, and Egger regression lines in green.

**Supplemental appendix 1:** OPCS-4 codes used to define the outcome “other revascularisation”

| Code | Description of other revascularisation |
|---|---|
| L311 | Percutaneous transluminal angioplasty of carotid artery |
| L313 | Endovascular repair of carotid artery |
| L314 | Percutaneous transluminal insertion of a stent into carotid artery |
| L318 | Other specified transluminal operations on carotid artery |
| L319 | Unspecified transluminal operations on carotid artery |
| L291 | Replacement of carotid artery using graft |
| L294 | Endarterectomy of carotid artery and patch repair of carotid artery |
| L295 | Endarterectomy of carotid artery NEC |
| L293 | Bypass to carotid artery NEC |
| L371 | Bypass of subclavian artery NEC |
| L372 | Endarterectomy of vertebral artery |
| L373 | Endarterectomy of subclavian artery and patch repair of subclavian artery |
| L374 | Endarterectomy of subclavian artery NEC |
| L378 | Other specified of subclavian artery |
| L379 | Unspecified reconstruction of subclavian artery |
| L383 | Open embolectomy of subclavian artery |
| L391 | Percutaneous transluminal angioplasty of subclavian artery |
| L392 | Percutaneous transluminal embolectomy of subclavian artery |
| L395 | Percutaneous transluminal insertion of stent into subclavian artery |
| L398 | Other specified transluminal operations on subclavian artery |
| L399 | Unspecified transluminal operations on subclavian artery |
| L411 | Plastic repair of renal artery and end to end anastomosis of renal artery |
| L412 | Bypass of renal artery |
| L413 | Replantation of renal artery |
| L414 | Endarterectomy of renal artery |
| L415 | Translocation of branch of renal artery |
| L416 | Patch angioplasty of renal artery |
| L418 | Other specified reconstruction of renal artery |
| L419 | Unspecified reconstruction of renal artery |
| L421 | Open embolectomy of renal artery |
| L422 | Open embolisation of renal artery |
| L424 | Operations on aneurysm of renal artery |
| L428 | Other specified other open operations on renal artery |
| L429 | Unspecified other open operations on renal artery |
| L431 | Percutaneous transluminal angioplasty of renal artery |
| L432 | Percutaneous transluminal embolectomy of renal artery |
| L435 | Percutaneous transluminal insertion of stent into renal artery |
| L438 | Other specified transluminal operations on renal artery |
| L439 | Unspecified transluminal operations on renal artery |
| L461 | Open embolectomy of visceral branch of abdominal aorta NEC |
| L471 | Percutaneous transluminal angioplasty of visceral branch of abdominal aorta |
| L474 | Percutaneous transluminal insertion of stent into visceral branch of abdominal aorta NEC |
| L478 | Other specified transluminal operations on other visceral branch of abdominal aorta |
| L479 | Unspecified transluminal operations on other visceral branch of abdominal aorta |
| L481 | Emergency replacement of aneurysmal common iliac artery by anastomosis of aorta to common iliac artery |
| L482 | Emergency replacement of aneurysmal iliac artery by anastomosis of aorta to external iliac artery |
| L483 | Emergency replacement of aneurysmal artery of leg by anastomosis of aorta to common femoral artery |
| L484 | Emergency replacement of aneurysmal artery of leg by anastomosis of aorta to superficial femoral artery |
| L485 | Emergency replacement of aneurysmal iliac artery by anastomosis of iliac artery to iliac artery |
| L486 | Emergency replacement of aneurysmal artery of leg by anastomosis of iliac artery to femoral artery |
| L488 | Other specified emergency replacement of aneurysmal iliac artery |
| L489 | Unspecified emergency replacement of aneurysmal iliac artery |
| L501 | Emergency bypass of common iliac artery by anastomosis of aorta to common iliac artery NEC |
| L502 | Emergency bypass of iliac artery by anastomosis of aorta to external iliac artery NEC |
| L503 | Emergency bypass of artery of leg by anastomosis of aorta to common femoral artery NEC |
| L504 | Emergency bypass of artery of leg by anastomosis of aorta to deep femoral artery NEC |
| L505 | Emergency bypass of iliac artery by anastomosis of iliac artery to iliac artery NEC |
| L506 | Emergency bypass of artery of leg by anastomosis of iliac artery to femoral artery NEC |
| L508 | Other specified other emergency bypass of iliac artery |
| L509 | Unspecified other emergency bypass of iliac artery |
| L511 | Bypass of common iliac artery by anastomosis of aorta to common iliac artery NEC |
| L512 | Bypass of iliac artery by anastomosis of aorta to external iliac artery NEC |
| L513 | Bypass of artery of leg by anastomosis of aorta to common femoral artery NEC |
| L514 | Bypass of artery of leg by anastomosis of aorta to deep femoral artery NEC |

| Code | Description of other revascularisation |
|---|---|
| L515 | Bypass of iliac artery by anastomosis of iliac artery to iliac artery NEC |
| L516 | Bypass of artery of leg by anastomosis of iliac artery to femoral artery NEC |
| L518 | Other specified other bypass of iliac artery |
| L519 | Unspecified other bypass of iliac artery |
| L521 | Endarterectomy of iliac artery and patch repair of iliac artery |
| L522 | Endarterectomy of iliac artery NEC |
| L528 | Other specified reconstruction of iliac artery |
| L529 | Unspecified reconstruction of iliac artery |
| L531 | Repair of iliac artery NEC |
| L532 | Open embolectomy of iliac artery |
| L533 | Operations on aneurysm of iliac artery NEC |
| L538 | Other specified open operations on iliac artery |
| L539 | Unspecified other open operations on iliac artery |
| L541 | Percutaneous transluminal angioplasty of iliac artery |
| L542 | Percutaneous transluminal embolectomy of iliac artery |
| L544 | Percutaneous transluminal insertion of stent into iliac artery |
| L548 | Other specified transluminal operations on iliac artery |
| L549 | Unspecified transluminal operations on iliac artery |
| L561 | Emergency replacement of aneurysmal femoral artery by anastomosis of femoral artery to femoral artery |
| L562 | Emergency replacement of aneurysmal femoral artery by anastomosis of femoral artery to popliteal artery using prosthesis |
| L563 | Emergency replacement of aneurysmal femoral artery by anastomosis of femoral artery to popliteal artery using vein graft |
| L564 | Emergency replacement of aneurysmal femoral artery by anastomosis of femoral artery to tibial artery using prosthesis |
| L565 | Emergency replacement of aneurysmal femoral artery by anastomosis of femoral artery to tibial artery using vein graft |
| L566 | Emergency replacement of aneurysmal femoral artery by anastomosis of femoral artery to peroneal artery using prosthesis |
| L567 | Emergency replacement of aneurysmal femoral artery by anastomosis of femoral artery to peroneal artery using vein graft |
| L568 | Other specified emergency replacement of aneurysmal femoral artery |
| L569 | Unspecified emergency replacement of aneurysmal femoral artery |
| L571 | Replacement of aneurysmal femoral artery by anastomosis of femoral artery to femoral artery NEC |
| L572 | Replacement of aneurysmal femoral artery by anastomosis of femoral artery to popliteal artery using prosthesis NEC |
| L573 | Replacement of aneurysmal femoral artery by anastomosis of femoral artery to popliteal artery using vein graft NEC |
| L574 | Replacement of aneurysmal femoral artery by anastomosis of femoral artery to tibial artery using prosthesis NEC |
| L575 | Replacement of aneurysmal femoral artery by anastomosis of femoral artery to tibial artery using vein graft NEC |
| L576 | Replacement of aneurysmal femoral artery by anastomosis of femoral artery to peroneal artery using prosthesis NEC |
| L577 | Replacement of aneurysmal femoral artery by anastomosis of femoral artery to peroneal artery using vein graft NEC |
| L578 | Other specified other replacement of aneurysmal femoral artery |
| L579 | Unspecified other replacement of aneurysmal femoral artery |
| L581 | Emergency bypass of femoral artery by anastomosis of femoral artery to femoral artery NEC |
| L582 | Emergency bypass of femoral artery by anastomosis of femoral artery to popliteal artery using prosthesis NEC |
| L583 | Emergency bypass of femoral artery by anastomosis of femoral artery to popliteal artery using vein graft NEC |
| L584 | Emergency bypass of femoral artery by anastomosis of femoral artery to tibial artery using prosthesis NEC |
| L585 | Emergency bypass of femoral artery by anastomosis of femoral artery to tibial artery using vein graft NEC |
| L586 | Emergency bypass of femoral artery by anastomosis of femoral artery to peroneal artery using prosthesis NEC |
| L587 | Emergency bypass of femoral artery by anastomosis of femoral artery to peroneal artery using vein graft NEC |
| L588 | Other specified other emergency bypass of femoral artery |
| L589 | Unspecified other emergency bypass of femoral artery |
| L591 | Bypass of femoral artery by anastomosis of femoral artery to femoral artery NEC |
| L592 | Bypass of femoral artery by anastomosis of femoral artery to popliteal artery using prosthesis NEC |
| L593 | Bypass of femoral artery by anastomosis of femoral artery to popliteal artery using vein graft NEC |
| L594 | Bypass of femoral artery by anastomosis of femoral artery to tibial artery using prosthesis NEC |
| L595 | Bypass of femoral artery by anastomosis of femoral artery to tibial artery using vein graft NEC |
| L596 | Bypass of femoral artery by anastomosis of femoral artery to peroneal artery using prosthesis NEC |
| L597 | Bypass of femoral artery by anastomosis of femoral artery to peroneal artery using vein graft NEC |
| L598 | Other specified other bypass of femoral artery |
| L599 | Unspecified other bypass of femoral artery |
| L601 | Endarterectomy of femoral artery and patch repair of femoral artery |
| L602 | Endarterectomy of femoral artery NEC |
| L603 | Profundoplasty of femoral artery and patch repair of deep femoral artery |
| L604 | Profundoplasty of femoral artery NEC |
| L608 | Other specified reconstruction of femoral artery |
| L609 | Unspecified reconstruction of femoral artery |
| L621 | Repair of femoral artery NEC |
| L622 | Repair of femoral artery NEC |
| L624 | Operations on aneurysm of femoral artery NEC |
| L628 | Other specified other open operations on femoral artery |

| Code | Description of other revascularisation |
|---|---|
| L629 | Unspecified other open operations on femoral artery |
| L631 | Percutaneous transluminal angioplasty of femoral artery |
| L632 | Percutaneous transluminal embolectomy of femoral artery |
| L633 | Percutaneous transluminal embolisation of femoral artery |
| L635 | Percutaneous transluminal insertion of stent into femoral artery |
| L638 | Other specified transluminal operations on femoral artery |
| L639 | Unspecified transluminal operations on femoral artery |
| L661 | Percutaneous transluminal arterial thrombolysis and reconstruction |
| L662 | Percutaneous transluminal stent reconstruction of artery |
| L665 | Percutaneous transluminal balloon angioplasty of artery |
| L667 | Percutaneous transluminal placement of peripheral stent in artery |
| L668 | Other specified other therapeutic transluminal operations on artery |
| L669 | Unspecified other therapeutic transluminal operations on artery |
| L718 | Other specified therapeutic transluminal operations on other artery |
| L719 | Unspecified therapeutic transluminal operations on other artery |
| L681 | Endarterectomy and patch repair of artery NEC |
| L682 | Endarterectomy NEC |

**Supplemental appendix 2:** ICD-10 diagnostic codes used to define hospitalisation for infection

| Code | Description of infection |
|---|---|
| A01 | Typhoid and paratyphoid fevers |
| A010 | Typhoid fever |
| A011 | Paratyphoid fever A |
| A012 | Paratyphoid fever B |
| A013 | Paratyphoid fever C |
| A014 | Paratyphoid fever, unspecified |
| A021 | Salmonella sepsis |
| A047 | Enterocolitis due to Clostridium difficile |
| A06 | Amebiasis |
| A060 | Acute amebic dysentery |
| A061 | Chronic intestinal amebiasis |
| A062 | Amebic nondysenteric colitis |
| A063 | Ameboma of intestine |
| A064 | Amebic liver abscess |
| A065 | Amebic lung abscess |
| A066 | Amebic brain abscess |
| A067 | Cutaneous amebiasis |
| A068 | Amebic infection of other sites |
| A069 | Amebiasis, unspecified |
| A15 | Respiratory tuberculosis |
| A150 | Tuberculosis of lung |
| A154 | Tuberculosis of intrathoracic lymph nodes |
| A155 | Tuberculosis of larynx, trachea and bronchus |
| A156 | Tuberculous pleurisy |
| A157 | Primary respiratory tuberculosis |
| A158 | Other respiratory tuberculosis |
| A159 | Respiratory tuberculosis unspecified |
| A17 | Tuberculosis of nervous system |
| A170 | Tuberculous meningitis |
| A171 | Meningeal tuberculoma |
| A178 | Other tuberculosis of nervous system |
| A179 | Tuberculosis of nervous system, unspecified |
| A18 | Tuberculosis of other organs |
| A180 | Tuberculosis of bones and joints |
| A181 | Tuberculosis of genitourinary system |
| A182 | Tuberculous peripheral lymphadenopathy |
| A183 | Tuberculosis of intestines, peritoneum and mesenteric glands |
| A184 | Tuberculosis of skin and subcutaneous tissue |
| A185 | Tuberculosis of eye |
| A186 | Tuberculosis of (inner) (middle) ear |
| A187 | Tuberculosis of adrenal glands |
| A188 | Tuberculosis of other specified organs |
| A19 | Miliary tuberculosis |
| A190 | Acute miliary tuberculosis of a single specified site |
| A191 | Acute miliary tuberculosis of multiple sites |
| A192 | Acute miliary tuberculosis, unspecified |
| A198 | Other miliary tuberculosis |
| A199 | Miliary tuberculosis, unspecified |
| A20 | Plague |
| A200 | Bubonic plague |
| A201 | Cellulocutaneous plague |
| A202 | Pneumonic plague |
| A203 | Plague meningitis |
| A207 | Septicemic plague |
| A208 | Other forms of plague |
| A209 | Plague, unspecified |
| A21 | Tularemia |
| A210 | Ulceroglandular tularemia |
| A211 | Oculoglandular tularemia |
| A212 | Pulmonary tularemia |
| A213 | Gastrointestinal tularemia |
| A217 | Generalized tularemia |
| A218 | Other forms of tularemia |
| A219 | Tularemia, unspecified |
| A22 | Anthrax |
| A220 | Cutaneous anthrax |

| Code | Description of infection |
|---|---|
| A221 | Pulmonary anthrax |
| A222 | Gastrointestinal anthrax |
| A227 | Anthrax sepsis |
| A228 | Other forms of anthrax |
| A229 | Anthrax, unspecified |
| A23 | Brucellosis |
| A230 | Brucellosis due to Brucella melitensis |
| A231 | Brucellosis due to Brucella abortus |
| A232 | Brucellosis due to Brucella suis |
| A233 | Brucellosis due to Brucella canis |
| A238 | Other brucellosis |
| A239 | Brucellosis, unspecified |
| A24 | Glanders and melioidosis |
| A240 | Glanders |
| A241 | Acute and fulminating melioidosis |
| A242 | Subacute and chronic melioidosis |
| A243 | Other melioidosis |
| A249 | Melioidosis, unspecified |
| A25 | Rat-bite fevers |
| A250 | Spirillosis |
| A251 | Streptobacillosis |
| A259 | Rat-bite fever, unspecified |
| A26 | Erysipeloid |
| A260 | Cutaneous erysipeloid |
| A267 | Erysipelothrix sepsis |
| A268 | Other forms of erysipeloid |
| A269 | Erysipeloid, unspecified |
| A27 | Leptospirosis |
| A270 | Leptospirosis icterohemorrhagica |
| A278 | Other forms of leptospirosis |
| A279 | Leptospirosis, unspecified |
| A28 | Other zoonotic bacterial diseases, not elsewhere classified |
| A280 | Pasteurellosis |
| A281 | Cat-scratch disease |
| A282 | Extraintestinal yersiniosis |
| A288 | Other specified zoonotic bacterial diseases, not elsewhere classified |
| A289 | Zoonotic bacterial disease, unspecified |
| A30 | Leprosy [Hansen's disease] |
| A300 | Indeterminate leprosy |
| A301 | Tuberculoid leprosy |
| A302 | Borderline tuberculoid leprosy |
| A303 | Borderline leprosy |
| A304 | Borderline lepromatous leprosy |
| A305 | Lepromatous leprosy |
| A308 | Other forms of leprosy |
| A309 | Leprosy, unspecified |
| A31 | Infection due to other mycobacteria |
| A310 | Pulmonary mycobacterial infection |
| A311 | Cutaneous mycobacterial infection |
| A312 | Disseminated mycobacterium avium-intracellulare complex (DMAC) |
| A318 | Other mycobacterial infections |
| A319 | Mycobacterial infection, unspecified |
| A32 | Listeriosis |
| A320 | Cutaneous listeriosis |
| A321 | Listerial meningitis and meningoencephalitis |
| A327 | Listerial sepsis |
| A328 | Other forms of listeriosis |
| A329 | Listeriosis, unspecified |
| A33 | Tetanus neonatorum |
| A34 | Obstetrical tetanus |
| A35 | Other tetanus |
| A36 | Diphtheria |
| A360 | Pharyngeal diphtheria |
| A361 | Nasopharyngeal diphtheria |
| A362 | Laryngeal diphtheria |
| A363 | Cutaneous diphtheria |
| A368 | Other diphtheria |

| Code | Description of infection |
|---|---|
| A369 | Diphtheria, unspecified |
| A37 | Whooping cough |
| A370 | Whooping cough due to Bordetella pertussis |
| A371 | Whooping cough due to Bordetella parapertussis |
| A378 | Whooping cough due to other Bordetella species |
| A379 | Whooping cough, unspecified species |
| A38 | Scarlet fever |
| A380 | Scarlet fever with otitis media |
| A381 | Scarlet fever with myocarditis |
| A388 | Scarlet fever with other complications |
| A389 | Scarlet fever, uncomplicated |
| A39 | Meningococcal infection |
| A390 | Meningococcal meningitis |
| A391 | Waterhouse-Friderichsen syndrome |
| A392 | Acute meningococcemia |
| A393 | Chronic meningococcemia |
| A394 | Meningococcemia, unspecified |
| A395 | Meningococcal heart disease |
| A398 | Other meningococcal infections |
| A399 | Meningococcal infection, unspecified |
| A40 | Streptococcal sepsis |
| A400 | Sepsis due to streptococcus, group A |
| A401 | Sepsis due to streptococcus, group B |
| A403 | Sepsis due to Streptococcus pneumoniae |
| A408 | Other streptococcal sepsis |
| A409 | Streptococcal sepsis, unspecified |
| A41 | Other sepsis |
| A410 | Sepsis due to Staphylococcus aureus |
| A411 | Sepsis due to other specified staphylococcus |
| A412 | Sepsis due to unspecified staphylococcus |
| A413 | Sepsis due to Hemophilus influenzae |
| A414 | Sepsis due to anaerobes |
| A415 | Sepsis due to other Gram-negative organisms |
| A418 | Other specified sepsis |
| A419 | Sepsis, unspecified organism |
| A42 | Actinomycosis |
| A420 | Pulmonary actinomycosis |
| A421 | Abdominal actinomycosis |
| A422 | Cervicofacial actinomycosis |
| A427 | Actinomycotic sepsis |
| A428 | Other forms of actinomycosis |
| A429 | Actinomycosis, unspecified |
| A43 | Nocardiosis |
| A430 | Pulmonary nocardiosis |
| A431 | Cutaneous nocardiosis |
| A438 | Other forms of nocardiosis |
| A439 | Nocardiosis, unspecified |
| A44 | Bartonellosis |
| A440 | Systemic bartonellosis |
| A441 | Cutaneous and mucocutaneous bartonellosis |
| A448 | Other forms of bartonellosis |
| A449 | Bartonellosis, unspecified |
| A46 | Erysipelas |
| A480 | Gas gangrene |
| A481 | Legionnaires' disease |
| A482 | Nonpneumonic Legionnaires' disease [Pontiac fever] |
| A483 | Toxic shock syndrome |
| A484 | Brazilian purpuric fever |
| A490 | Staphylococcal infection, unspecified site |
| A491 | Streptococcal infection, unspecified site |
| A492 | Hemophilus influenzae infection, unspecified site |
| A493 | Mycoplasma infection, unspecified site |
| A50 | Congenital syphilis |
| A500 | Early congenital syphilis, symptomatic |
| A501 | Early congenital syphilis, latent |
| A502 | Early congenital syphilis, unspecified |
| A503 | Late congenital syphilitic oculopathy |

| Code | Description of infection |
|---|---|
| A504 | Late congenital neurosyphilis [juvenile neurosyphilis] |
| A505 | Other late congenital syphilis, symptomatic |
| A506 | Late congenital syphilis, latent |
| A507 | Late congenital syphilis, unspecified |
| A509 | Congenital syphilis, unspecified |
| A51 | Early syphilis |
| A510 | Primary genital syphilis |
| A511 | Primary anal syphilis |
| A512 | Primary syphilis of other sites |
| A513 | Secondary syphilis of skin and mucous membranes |
| A514 | Other secondary syphilis |
| A515 | Early syphilis, latent |
| A519 | Early syphilis, unspecified |
| A52 | Late syphilis |
| A520 | Cardiovascular and cerebrovascular syphilis |
| A521 | Symptomatic neurosyphilis |
| A522 | Asymptomatic neurosyphilis |
| A523 | Neurosyphilis, unspecified |
| A527 | Other symptomatic late syphilis |
| A528 | Late syphilis, latent |
| A529 | Late syphilis, unspecified |
| A53 | Other and unspecified syphilis |
| A530 | Latent syphilis, unspecified as early or late |
| A539 | Syphilis, unspecified |
| A54 | Gonococcal infection |
| A540 | Gonococcal infection of lower genitourinary tract without periurethral or accessory gland abscess |
| A541 | Gonococcal infection of lower genitourinary tract with periurethral and accessory gland abscess |
| A542 | Gonococcal pelviperitonitis and other gonococcal genitourinary infection |
| A543 | Gonococcal infection of eye |
| A544 | Gonococcal infection of musculoskeletal system |
| A545 | Gonococcal pharyngitis |
| A546 | Gonococcal infection of anus and rectum |
| A548 | Other gonococcal infections |
| A549 | Gonococcal infection, unspecified |
| A55 | Chlamydial lymphogranuloma (venereum) |
| A56 | Other sexually transmitted chlamydial diseases |
| A560 | Chlamydial infection of lower genitourinary tract |
| A561 | Chlamydial infection of pelviperitoneum and other genitourinary organs |
| A562 | Chlamydial infection of genitourinary tract, unspecified |
| A563 | Chlamydial infection of anus and rectum |
| A564 | Chlamydial infection of pharynx |
| A568 | Sexually transmitted chlamydial infection of other sites |
| A57 | Chancroid |
| A58 | Granuloma inguinale |
| A59 | Trichomoniasis |
| A590 | Urogenital trichomoniasis |
| A598 | Trichomoniasis of other sites |
| A599 | Trichomoniasis, unspecified |
| A65 | Nonvenereal syphilis |
| A66 | Yaws |
| A660 | Initial lesions of yaws |
| A661 | Multiple papillomata and wet crab yaws |
| A662 | Other early skin lesions of yaws |
| A663 | Hyperkeratosis of yaws |
| A664 | Gummata and ulcers of yaws |
| A665 | Gangosa |
| A666 | Bone and joint lesions of yaws |
| A667 | Other manifestations of yaws |
| A668 | Latent yaws |
| A669 | Yaws, unspecified |
| A67 | Pinta [carate] |
| A670 | Primary lesions of pinta |
| A671 | Intermediate lesions of pinta |
| A672 | Late lesions of pinta |
| A673 | Mixed lesions of pinta |
| A679 | Pinta, unspecified |
| A68 | Relapsing fevers |

| Code | Description of infection |
|---|---|
| A680 | Louse-borne relapsing fever |
| A681 | Tick-borne relapsing fever |
| A689 | Relapsing fever, unspecified |
| A69 | Other spirochetal infections |
| A690 | Necrotizing ulcerative stomatitis |
| A691 | Other Vincent's infections |
| A692 | Lyme disease |
| A698 | Other specified spirochetal infections |
| A699 | Spirochetal infection, unspecified |
| A70 | Chlamydia psittaci infections |
| A71 | Trachoma |
| A710 | Initial stage of trachoma |
| A711 | Active stage of trachoma |
| A719 | Trachoma, unspecified |
| A74 | Other diseases caused by chlamydiae |
| A740 | Chlamydial conjunctivitis |
| A748 | Other chlamydial diseases |
| A749 | Chlamydial infection, unspecified |
| A75 | Typhus fever |
| A750 | Epidemic louse-borne typhus fever due to Rickettsia prowazekii |
| A751 | Recrudescent typhus [Brill's disease] |
| A752 | Typhus fever due to Rickettsia typhi |
| A753 | Typhus fever due to Rickettsia tsutsugamushi |
| A759 | Typhus fever, unspecified |
| A77 | Spotted fever [tick-borne rickettsioses] |
| A770 | Spotted fever due to Rickettsia rickettsii |
| A771 | Spotted fever due to Rickettsia conorii |
| A772 | Spotted fever due to Rickettsia siberica |
| A773 | Spotted fever due to Rickettsia australis |
| A774 | Ehrlichiosis |
| A778 | Other spotted fevers |
| A779 | Spotted fever, unspecified |
| A78 | Q fever |
| A79 | Other rickettsioses |
| A790 | Trench fever |
| A791 | Rickettsialpox due to Rickettsia akari |
| A798 | Other specified rickettsioses |
| A799 | Rickettsiosis, unspecified |
| B58 | Toxoplasmosis |
| B580 | Toxoplasma oculopathy |
| B581 | Toxoplasma hepatitis |
| B582 | Toxoplasma meningoencephalitis |
| B583 | Pulmonary toxoplasmosis |
| B588 | Toxoplasmosis with other organ involvement |
| B589 | Toxoplasmosis, unspecified |
| B59 | Pneumocystosis |
| B600 | Babesiosis |
| B601 | Acanthamebiasis |
| B602 | Naegleriasis |
| B90 | Sequelae of tuberculosis |
| B900 | Sequelae of central nervous system tuberculosis |
| B901 | Sequelae of genitourinary tuberculosis |
| B902 | Sequelae of tuberculosis of bones and joints |
| B908 | Sequelae of tuberculosis of other organs |
| B909 | Sequelae of respiratory and unspecified tuberculosis |
| B92 | Sequelae of leprosy |
| B940 | Sequelae of trachoma |
| B95 | Streptococcus, Staphylococcus, and Enterococcus as the cause of diseases classified elsewhere |
| B950 | Streptococcus, group A, as the cause of diseases classified elsewhere |
| B951 | Streptococcus, group B, as the cause of diseases classified elsewhere |
| B952 | Enterococcus as the cause of diseases classified elsewhere |
| B953 | Streptococcus pneumoniae as the cause of diseases classified elsewhere |
| B954 | Other streptococcus as the cause of diseases classified elsewhere |
| B955 | Unspecified streptococcus as the cause of diseases classified elsewhere |
| B956 | Staphylococcus aureus as the cause of diseases classified elsewhere |
| B957 | Other staphylococcus as the cause of diseases classified elsewhere |
| B958 | Unspecified staphylococcus as the cause of diseases classified elsewhere |

| Code | Description of infection |
|---|---|
| B960 | Mycoplasma pneumoniae [M. pneumoniae] as the cause of diseases classified elsewhere |
| B961 | Klebsiella pneumoniae [K. pneumoniae] as the cause of diseases classified elsewhere |
| B963 | Hemophilus influenzae [H. influenzae] as the cause of diseases classified elsewhere |
| B964 | Proteus (mirabilis) (morganii) as the cause of diseases classified elsewhere |
| B965 | Pseudomonas (aeruginosa) (mallei) (pseudomallei) as the cause of diseases classified elsewhere |
| B966 | Bacteroides fragilis [B. fragilis] as the cause of diseases classified elsewhere |
| B967 | Clostridium perfringens [C. perfringens] as the cause of diseases classified elsewhere |
| B968 | Other specified bacterial agents as the cause of diseases classified elsewhere |
| D733 | Abscess of spleen |
| E321 | Abscess of thymus |
| G00 | Bacterial meningitis, not elsewhere classified |
| G000 | Hemophilus meningitis |
| G001 | Pneumococcal meningitis |
| G002 | Streptococcal meningitis |
| G003 | Staphylococcal meningitis |
| G008 | Other bacterial meningitis |
| G009 | Bacterial meningitis, unspecified |
| G01 | Meningitis in bacterial diseases classified elsewhere |
| G042 | Bacterial meningoencephalitis and meningomyelitis, not elsewhere classified |
| G06 | Intracranial and intraspinal abscess and granuloma |
| G060 | Intracranial abscess and granuloma |
| G061 | Intraspinal abscess and granuloma |
| G062 | Extradural and subdural abscess, unspecified |
| G07 | Intracranial and intraspinal abscess and granuloma in diseases classified elsewhere |
| H050 | Acute inflammation of orbit |
| H602 | Malignant otitis externa |
| H700 | Acute mastoiditis |
| H701 | Chronic mastoiditis |
| H702 | Petrositis |
| H708 | Other mastoiditis and related conditions |
| H709 | Unspecified mastoiditis |
| H750 | Mastoiditis in infectious and parasitic diseases classified elsewhere |
| I00 | Rheumatic fever without heart involvement |
| I01 | Rheumatic fever with heart involvement |
| I010 | Acute rheumatic pericarditis |
| I011 | Acute rheumatic endocarditis |
| I012 | Acute rheumatic myocarditis |
| I018 | Other acute rheumatic heart disease |
| I019 | Acute rheumatic heart disease, unspecified |
| I02 | Rheumatic chorea |
| I020 | Rheumatic chorea with heart involvement |
| I029 | Rheumatic chorea without heart involvement |
| I33 | Acute and subacute endocarditis |
| I330 | Acute and subacute infective endocarditis |
| I339 | Acute and subacute endocarditis, unspecified |
| I38 | Endocarditis, valve unspecified |
| I39 | Endocarditis and heart valve disorders in diseases classified elsewhere |
| I76 | Septic arterial embolism |
| I96 | Gangrene, not elsewhere classified |
| J020 | Streptococcal pharyngitis |
| J030 | Streptococcal tonsillitis |
| J13 | Pneumonia due to Streptococcus pneumoniae |
| J14 | Pneumonia due to Hemophilus influenzae |
| J15 | Bacterial pneumonia, not elsewhere classified |
| J150 | Pneumonia due to Klebsiella pneumoniae |
| J151 | Pneumonia due to Pseudomonas |
| J152 | Pneumonia due to staphylococcus |
| J153 | Pneumonia due to streptococcus, group B |
| J154 | Pneumonia due to other streptococci |
| J155 | Pneumonia due to Escherichia coli |
| J156 | Pneumonia due to other Gram-negative bacteria |
| J157 | Pneumonia due to Mycoplasma pneumoniae |
| J158 | Pneumonia due to other specified bacteria |
| J159 | Unspecified bacterial pneumonia |
| J16 | Pneumonia due to other infectious organisms, not elsewhere classified |
| J160 | Chlamydial pneumonia |
| J168 | Pneumonia due to other specified infectious organisms |

| Code | Description of infection |
|---|---|
| J17 | Pneumonia in diseases classified elsewhere |
| J18 | Pneumonia, unspecified organism |
| J180 | Bronchopneumonia, unspecified organism |
| J181 | Lobar pneumonia, unspecified organism |
| J182 | Hypostatic pneumonia, unspecified organism |
| J188 | Other pneumonia, unspecified organism |
| J189 | Pneumonia, unspecified organism |
| J36 | Peritonsillar abscess |
| J390 | Retropharyngeal and parapharyngeal abscess |
| J391 | Other abscess of pharynx |
| J85 | Abscess of lung and mediastinum |
| J850 | Gangrene and necrosis of lung |
| J851 | Abscess of lung with pneumonia |
| J852 | Abscess of lung without pneumonia |
| J853 | Abscess of mediastinum |
| J86 | Pyothorax |
| J860 | Pyothorax with fistula |
| J869 | Pyothorax without fistula |
| K046 | Periapical abscess with sinus |
| K047 | Periapical abscess without sinus |
| K113 | Abscess of salivary gland |
| K122 | Cellulitis and abscess of mouth |
| K35 | Acute appendicitis |
| K352 | Acute appendicitis with generalized peritonitis |
| K353 | Acute appendicitis with localized peritonitis |
| K358 | Other and unspecified acute appendicitis |
| K36 | Other appendicitis |
| K37 | Unspecified appendicitis |
| K401 | Bilateral inguinal hernia, with gangrene |
| K404 | Unilateral inguinal hernia, with gangrene |
| K411 | Bilateral femoral hernia, with gangrene |
| K414 | Unilateral femoral hernia, with gangrene |
| K421 | Umbilical hernia with gangrene |
| K431 | Incisional hernia with gangrene |
| K434 | Parastomal hernia with gangrene |
| K437 | Other and unspecified ventral hernia with gangrene |
| K441 | Diaphragmatic hernia with gangrene |
| K451 | Other specified abdominal hernia with gangrene |
| K461 | Unspecified abdominal hernia with gangrene |
| K570 | Diverticulitis of small intestine with perforation and abscess |
| K572 | Diverticulitis of large intestine with perforation and abscess |
| K574 | Diverticulitis of both small and large intestine with perforation and abscess |
| K578 | Diverticulitis of intestine, part unspecified, with perforation and abscess |
| K630 | Abscess of intestine |
| K650 | Generalized (acute) peritonitis |
| K651 | Peritoneal abscess |
| K652 | Spontaneous bacterial peritonitis |
| K653 | Choleperitonitis |
| K658 | Other peritonitis |
| K659 | Peritonitis, unspecified |
| K681 | Retroperitoneal abscess |
| K750 | Abscess of liver |
| K800 | Calculus of gallbladder with acute cholecystitis |
| K801 | Calculus of gallbladder with other cholecystitis |
| K803 | Calculus of bile duct with cholangitis |
| K804 | Calculus of bile duct with cholecystitis |
| K806 | Calculus of gallbladder and bile duct with cholecystitis |
| K81 | Cholecystitis |
| K810 | Acute cholecystitis |
| K811 | Chronic cholecystitis |
| K812 | Acute cholecystitis with chronic cholecystitis |
| K819 | Cholecystitis, unspecified |
| K830 | Cholangitis |
| K901 | Tropical sprue |
| L00 | Staphylococcal scalded skin syndrome |
| M00 | Pyogenic arthritis |
| M000 | Staphylococcal arthritis and polyarthritis |

| Code | Description of infection |
|---|---|
| M001 | Pneumococcal arthritis and polyarthritis |
| M002 | Other streptococcal arthritis and polyarthritis |
| M008 | Arthritis and polyarthritis due to other bacteria |
| M009 | Pyogenic arthritis, unspecified |
| M01 | Direct infections of joint in infectious and parasitic diseases classified elsewhere |
| M01X | Direct infection of joint in infectious and parasitic diseases classified elsewhere |
| M462 | Osteomyelitis of vertebra |
| M463 | Infection of intervertebral disc (pyogenic) |
| M650 | Abscess of tendon sheath |
| M651 | Other infective (teno)synovitis |
| M726 | Necrotizing fasciitis |
| M86 | Osteomyelitis |
| M860 | Acute hematogenous osteomyelitis |
| M861 | Other acute osteomyelitis |
| M862 | Subacute osteomyelitis |
| M863 | Chronic multifocal osteomyelitis |
| M864 | Chronic osteomyelitis with draining sinus |
| M865 | Other chronic hematogenous osteomyelitis |
| M866 | Other chronic osteomyelitis |
| M868 | Other osteomyelitis |
| M869 | Osteomyelitis, unspecified |
| N10 | Acute pyelonephritis |
| N151 | Renal and perinephric abscess |
| N300 | Acute cystitis |
| N303 | Trigonitis |
| N340 | Urethral abscess |
| N390 | Urinary tract infection, site not specified |
| N410 | Acute prostatitis |
| N412 | Abscess of prostate |
| N431 | Infected hydrocele |
| N454 | Abscess of epididymis or testis |
| N493 | Fournier gangrene |
| N70 | Salpingitis and oophoritis |
| N700 | Acute salpingitis and oophoritis |
| N701 | Chronic salpingitis and oophoritis |
| N709 | Salpingitis and oophoritis, unspecified |
| N71 | Inflammatory disease of uterus, except cervix |
| N710 | Acute inflammatory disease of uterus |
| N711 | Chronic inflammatory disease of uterus |
| N719 | Inflammatory disease of uterus, unspecified |
| N72 | Inflammatory disease of cervix uteri |
| N730 | Acute parametritis and pelvic cellulitis |
| N731 | Chronic parametritis and pelvic cellulitis |
| N732 | Unspecified parametritis and pelvic cellulitis |
| N733 | Female acute pelvic peritonitis |
| N734 | Female chronic pelvic peritonitis |
| N735 | Female pelvic peritonitis, unspecified |
| N738 | Other specified female pelvic inflammatory diseases |
| N739 | Female pelvic inflammatory disease, unspecified |
| N74 | Female pelvic inflammatory disorders in diseases classified elsewhere |
| N980 | Infection associated with artificial insemination |
| T802 | Infections following infusion, transfusion and therapeutic injection |
| T826 | Infection and inflammatory reaction due to cardiac valve prosthesis |
| T827 | Infection and inflammatory reaction due to other cardiac and vascular devices, implants and grafts |
| T835 | Infection and inflammatory reaction due to prosthetic device, implant and graft in urinary system |
| T836 | Infection and inflammatory reaction due to prosthetic device, implant and graft in genital tract |
| T845 | Infection and inflammatory reaction due to internal joint prosthesis |
| T846 | Infection and inflammatory reaction due to internal fixation device |
| T847 | Infection and inflammatory reaction due to other internal orthopedic prosthetic devices, implants and grafts |
| T857 | Infection and inflammatory reaction due to other internal prosthetic devices, implants and grafts |
| T874 | Infection of amputation stump |
| T875 | Necrosis of amputation stump |
